# Germinal center responses to SARS-CoV-2 mRNA vaccines in healthy and immunocompromised individuals

**DOI:** 10.1101/2021.09.16.21263686

**Authors:** Katlyn Lederer, Kalpana Parvathaneni, Mark M. Painter, Emily Bettini, Divyansh Agarwal, Kendall A. Lundgreen, Madison Weirick, Rishi R. Goel, Xiaoming Xu, Elizabeth M. Drapeau, Sigrid Gouma, Allison R. Greenplate, Carole Le Coz, Neil Romberg, Lisa Jones, Mark Rosen, Behdad Besharatian, Mary Kaminiski, Daniela Weiskopf, Alessandro Sette, Scott E. Hensley, Paul Bates, E. John Wherry, Ali Naji, Vijay Bhoj, Michela Locci

## Abstract

Vaccine-mediated immunity often relies on the generation of protective antibodies and memory B cells, which commonly stem from germinal center (GC) reactions. An in-depth comparison of the GC responses elicited by SARS-CoV-2 mRNA vaccines in healthy and immunocompromised individuals has not yet been performed due to the challenge of directly probing human lymph nodes. In this study, through a fine-needle-aspiration-based approach, we profiled the immune responses to SARS-CoV-2 mRNA vaccines in lymph nodes of healthy individuals and kidney transplant (KTX) recipients. We found that, unlike healthy subjects, KTX recipients presented deeply blunted SARS-CoV-2-specific GC B cell responses coupled with severely hindered T follicular helper cells, SARS-CoV-2 receptor-binding-domain-specific memory B cells and neutralizing antibodies. KTX recipients also displayed reduced SARS-CoV-2-specific CD4 and CD8 T cell frequencies. Broadly, these data indicate impaired GC-derived immunity in immunocompromised individuals, and suggest a GC-origin for certain humoral and memory B cell responses following mRNA vaccination.

## INTRODUCTION

Messenger RNA (mRNA) vaccines have been intensively investigated over the past decade and shown to successfully induce long-lasting, protective immune responses in animal models (Awasthi et al., 2019; Espeseth et al., 2020; Freyn et al., 2020; Pardi et al., 2017, 2018b, 2018a; Richner et al., 2017). This vaccine platform was licensed for human use for the first time during the pandemic caused by severe acute respiratory syndrome coronavirus 2 (SARS-CoV-2) (Bettini and Locci, 2021; Carvalho et al., 2021; Krammer, 2020), and much still needs to be learned about the quality of the immune responses elicited by mRNA vaccines.

Most vaccines confer protection by eliciting antigen-specific antibodies (Abs) and memory B cells (Plotkin, 2010; Sallusto et al., 2010). Abs are secreted by plasma cells and constitute a critical immune endpoint of vaccination, as they can potentially neutralize pathogens and prevent infections. Equally important are memory B cells that act as a second line of defense and rapidly give rise to a quick burst of Ab-secreting plasma cells if the pathogens break through the “protective wall” of the pre-existing Abs. Plasma cells and memory B cells are commonly generated during germinal center (GC) reactions (Allen et al., 2007; Mesin et al., 2016) in vaccine-draining lymph nodes. In GCs, pathogen-activated B cells first undergo mutations in their immunoglobulin genes. Next, the high-affinity GC B cell clones resulting from this somatic hypermutation (SHM) process are positively selected, and ultimately differentiate into long-lived plasma cells and memory B cells. GC reactions are orchestrated by T follicular helper (Tfh) cells, specialized CD4 T cells that deliver a variety of signals shaping the fate of GC B cells (Crotty, 2019; Vinuesa et al., 2016). We and others have previously demonstrated that, in mice, mRNA vaccines can elicit potent GC responses that were closely intertwined with an efficient induction of SARS-CoV-2-specific binding-Abs, neutralizing (nAbs), and memory B cells (Lederer et al., 2020; Tai et al., 2020; Vogel et al., 2021). These data suggest that GC reactions might be crucial for the formation of durable nAb and memory B cells following SARS-CoV-2 vaccination. In line with these animal data, several studies have characterized the immune responses to the SARS-CoV-2 mRNA vaccines in humans and found a robust induction of nAbs and memory B cells (Bettini and Locci, 2021; Collier et al., 2021; Edara et al., 2021; Goel et al., 2021; Jackson et al., 2020; Planas et al., 2021; Sahin et al., 2020; Stamatatos et al., 2021; Walsh et al., 2020; Wang et al., 2021; Widge et al., 2020). However, with only one exception (Turner et al., 2021), all published human vaccine studies focused on the analysis of the immune responses measurable in peripheral blood. Hence, a deep evaluation of the GC reactions driven by SARS-CoV-2 mRNA vaccines in human, including their connection with nAbs and memory B cells, is still missing.

Another major question that warrants further investigation is whether SARS-CoV-2 mRNA vaccines can successfully promote high-quality immune responses in individuals lacking a fully functional immune system. Analyses of blood samples from recipients of solid organ transplants (SOT) who underwent SARS-CoV-2 vaccination yielded mixed results (Benotmane et al., 2021; Boyarsky et al., 2021a, 2021b; Cucchiari et al., 2021; Kamar et al., 2021; Massa et al., 2021; Rincon-Arevalo et al., 2021). Some of these studies suggested that a large fraction of SOT recipients can still generate detectable SARS-CoV-2 binding Ab titers, whereas others indicate that SOT recipients completely fail to produce B cell responses and Abs to SARS-CoV-2 mRNA vaccines. A common denominator, however, is the heavily curtailed nAb production in SOT recipients following immunization. While the evidence from studies conducted with blood samples hints at crippled GC formation, the exploration of GC responses to SARS-CoV-2 mRNA vaccination in SOT recipients still remains an uncharted territory.

Herein, by deploying a fine-needle aspiration (FNA) approach (Havenar-Daughton et al., 2020), we evaluated the GC responses elicited by SARS-CoV-2 mRNA vaccines in draining lymph nodes of healthy donors (HDs) and KTX recipients and assessed their connection to humoral and memory B cell responses. Our study uncovered a potent elicitation of SARS-CoV-2 full-length spike (Full S) and receptor binding domain (RBD)-specific GC B cells localized in vaccine-draining lymph nodes upon primary immunization of healthy individuals, which was further enhanced by the booster vaccination. Furthermore, SARS-CoV-2-specific GC B cell responses were associated with a robust induction of Tfh cells, RBD-specific memory B cells and nAbs. These finding were in stark contrast to a profound impairment of the GC responses in KTX recipients, which were coupled to a nearly-abolished RBD-specific memory B cell response and nAb formation and opposed to a measurable generation of S-specific memory B cells binding Full S outside the RBD region. Overall, this study shows that, in individuals with an intact immune system, RBD-specific memory B cells and nAbs are efficiently induced by SARS-CoV-2 mRNA vaccination and might have a GC origin. Conversely, these responses are not efficiently generated following vaccination in individuals receiving immunosuppressant drugs. This study has important implications for guiding future studies aimed at unraveling human immune responses after vaccination and for supporting the decision to perform additional booster immunizations against SARS-CoV-2 in people with a compromised immune system.

## RESULTS

### Robust SARS-CoV-2-specific GC B cell responses are elicited by mRNA vaccines and localized in draining lymph nodes of immunocompetent individuals

GC B cells and Tfh cells induced by vaccination/infection are only present in lymphoid tissues and cannot be studied in blood (Vella et al., 2019). To address this issue, we conducted a human study where lymphoid tissue immune responses elicited by SARS-CoV-2 mRNA vaccines were probed in healthy individuals via fine-needle aspiration (FNA). The FNA approach has been successfully used to track GC B cell responses to vaccination in humans, and is a relatively simple procedure carrying minimal risk that does not alter the architecture of lymph nodes (Havenar-Daughton et al., 2020; Turner et al., 2020, 2021). 15 healthy subjects (23-76 years old) were enrolled in this study prior to vaccination with BNT162b2 or mRNA-1273 (Table 1). FNA samples were collected two weeks after the first immunization (V2: day 14 +/- 2) and eight days after the booster immunization (V3: day 8 +/- 2) (Figure 1A). Matched blood samples were also obtained at the same time points and before vaccination (V1). Axillary draining lymph nodes from the same arm where the vaccines were administered (ipsilateral) were visualized by ultrasound to guide the FNA procedure (Figure 1B and Supplementary Video 1). In healthy subjects, the number of live cells recovered from the FNA procedure ranged from 0.3 to 40 x 10^6^ cells. A 23-parameter flow cytometry assay was performed on FNA samples to profile the immune responses induced by vaccination. GC B cells were defined as class-switched B cells co-expressing CD38, low-intermediate levels of CD27 and the signature transcription factor BCL6 (Figure 1C and Figure S1A). Pre-pandemic tonsil samples, which are highly enriched in GCs, and putative quiescent cadaveric lymph nodes from SARS-CoV-2 negative individuals were used as positive and negative controls, respectively. SARS-CoV-2 mRNA vaccination elicited detectable GC B cell responses after primary immunization, which were further enhanced by the booster vaccination (Figure 1C). The increase in GC B cell frequencies was measurable when the responses were evaluated following a longitudinal (11 individuals) or an orthogonal (15 individuals) approach (Figures 1D and S1B). No correlation between age and GC B cell frequencies was observed (Figure S1C). Next, by using a combination of fluorescently-labelled SARS-CoV-2 Full S and RBD tetrameric probes, we identified SARS-CoV-2-specific GC B cells as GC B cells binding the Full S but not the RBD probes (Full S^+^RBD^-^) or simultaneously binding the Full S and RBD probes (Full S^+^ RBD^+^), while failing to bind an irrelevant tetrameric probe (influenza hemagglutinin, HA from A/Puerto Rico/8/34) (Figure 1E and Figure S1A). The specificity of the probes is indicated by the lack of Full S and RBD-specific GC B cells in pre-pandemic tonsil samples (Figure 1E). Overall, a boost in both Full S RBD^-^ and RBD^+^ GC B cell frequencies following the second vaccine dose was observed, especially when a longitudinal evaluation was performed (Figure 1F and S1D). Similar to total GC B cells, SARS-CoV-2-specific GC B cells did not correlate with age (Figure S1E-F). Next, to determine if mRNA vaccine-induced GC responses were detectable in non-draining lymph nodes, we collected contralateral axillary lymph nodes, which do not directly drain the mRNA vaccines from the injection site, from a few vaccinees after the booster immunization (n=4). When compared to the matched ipsilateral lymph nodes, the contralateral lymph nodes displayed a trend for lower frequencies of GC B cells (Figure 1G-H), which were not SARS-CoV-2 specific (Figure 1I-J). Overall, these data demonstrate that, in immunocompetent subjects, SARS-CoV-2 mRNA vaccines efficiently elicit antigen-specific GC B cell responses that are enhanced by a booster immunization and localized in the ipsilateral axillary draining lymph nodes.

**Figure 1.**
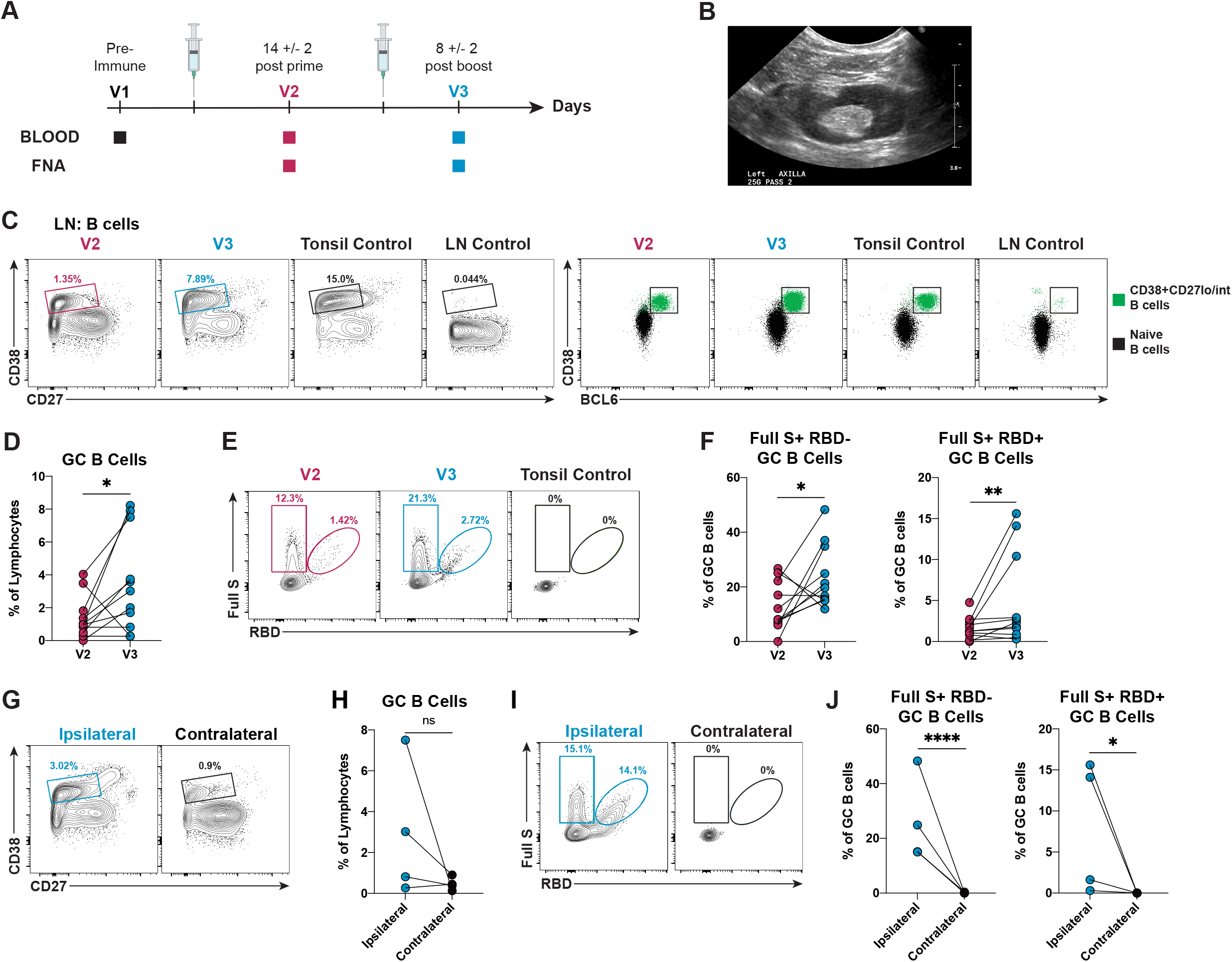
GC B cell responses to SARS-CoV-2 mRNA vaccines are detected in ipsilateral but not in contralateral lymph nodes of immunocompetent individuals. **(A)** Schematic of study design. Fifteen healthy donors (HD) and thirteen kidney transplant (KTX) recipients volunteered to receive two-doses of either BNT162b2 or mRNA-1273. Blood was collected before immunization (V1) and at approximately 14 days post-prime (V2) and 8 days post-boost (V3). FNAs were collected at approximately 14 days post-prime (V2) and 8 days post-boost (V3). **(B)** Representative ultrasound visualization of a HD axillary lymph node probed for the FNA procedure during V3 collection time point. **(C) (Left)** Representative flow cytometry plots of CD38^+^CD27^lo/int^ B cells in HD FNAs at V2 and V3, a pre-pandemic tonsil sample (Tonsil Control) and a putative quiescent cadaveric lymph node (LN Control). **(Right)** Representative flow cytometry plots of GC B cells (CD19^+^CD4^-^CD8^-^IgM^-^ IgD^-^CD38^+^CD27^lo/int^BCL6^+^) from the indicated donors. Naive B cells were used as negative control population to set the BCL6 gate. **(D)** Quantification of GC B cells in FNAs from HDs, displayed as a percentage of total lymphocytes. Paired (longitudinal) changes between V2 and V3 are displayed. **(E)** Representative flow cytometry of SARS-CoV-2 Full S^+^ RBD^-^ (CD19^+^CD4^-^CD8^-^IgM^-^IgD^-^ CD38^+^CD27^lo/int^BCL6^+^HA^-^Full S^+^RBD^-^) and Full S^+^ RBD^+^ (CD19^+^CD4^-^CD8^-^IgM^-^IgD^-^ CD38^+^CD27^lo/int^BCL6^+^HA^-^Full S^+^RBD^+^) GC B cells in HD FNAs at V2 and V3 and in a tonsil control. **(F)** Quantification of Full S^+^ RBD^-^ (**left**) and Full S^+^ RBD^+^ (**right**) GC B cells from HDs at V2 and V3, displayed as a percentage of GC B cells. **(G)** Representative flow cytometry plots of GC B cells in draining (ipsilateral) and in non-draining (contralateral) lymph nodes of HDs. **(H)** Quantification of GC B cells in FNAs at V3 from HD ipsilateral and contralateral lymph nodes, displayed as a percentage of total lymphocytes. **(I)** Representative flow cytometry plots of antigen-specific GC B cells in ipsilateral and contralateral lymph nodes at V3. **(J)** Quantification of Full S^+^ RBD^-^ (**left**) and Full S^+^ RBD^+^ (**right**) GC B cells in ipsilateral and contralateral lymph nodes, displayed as a percentage of GC B cells. In (D and F), n = 11 donors; red data points = V2 and blue data points = V3. In (H and J), n=4 donors; blue data points = ipsilateral lymph node and black data points = contralateral lymph node, both at V3. Statistical analysis: In (D and F), a paired Mann-Whitney U test with continuity correction was performed. In (H and J), the Wald-Wolfowitz runs test was performed. * P ≤ 0.05, ** P ≤ 0.01, *** P ≤ 0.001, **** P ≤ 0.0001.

### Tfh responses are detectable in draining lymph nodes of immunocompetent individuals upon SARS-CoV-2 mRNA vaccination

Tfh cells are CD4 T cells specialized in regulating GC responses. By enabling the selection of high affinity GC B cells and curbing the magnitude of GC reactions, Tfh cells modulate affinity maturation in infection and vaccination (Crotty, 2019). We measured the frequency of GC Tfh cells (referred to as Tfh cells) defined by the signature markers CXCR5 and PD-1 (Figures 2A and S2A). Expression of the lineage-defining transcription factor BCL6 confirmed the identity of this CXCR5^hi^PD-1^hi^ cell population as Tfh cells (Figure 2B). As anticipated, negligible Tfh cell frequencies were found in putative quiescent lymph nodes from cadaveric donors (Figure S2B) and the contralateral lymph nodes of vaccinees (Figure 2A), in contrast to a more abundant Tfh cell presence in tonsils (Figure S2B). Of note, the frequencies of Tfh cells in draining lymph nodes of vaccinated healthy subjects had a trend for higher values than in contralateral lymph nodes (p=0.056, Figure 2C) and increased after the second vaccine dose (Figures 2D and S2C). Tfh cells are a functionally heterogenous population that, in humans, is often functionally stratified by chemokine receptor expression based on extensive work performed on human circulating Tfh cells (Ueno, 2016). CXCR3-expressing Tfh cells are Th1-polarized (Locci et al., 2013; Morita et al., 2011). By contrast, CXCR3^-^ Tfh cells can be distinguished by CCR6 expression into Th2 (CCR6^-^) and Th17 (CCR6^+^)-polarized cells (Morita et al., 2011). CCR4 was also used in this analysis to help refine the delineation of Th2 (CCR6^-^CCR4^+^) and Th17 (CCR6^+^CCR4^+^)-biased cells (Figure 2E) (Acosta-Rodriguez et al., 2007). This analysis approach showed that the Tfh cells present in the draining lymph nodes of vaccinees comprised Th1 and Th2-polarized Tfh cells, but not Th17-biased Tfh cells (Figure 2F). Of note, Tfh cells present in vaccine draining lymph nodes correlated with both Full S^+^ RBD^-^ GC B cells and RBD-specific GC B cells (Figure 2G). While bona fide Tfh cells can be found exclusively in secondary lymphoid organs (Vella et al., 2019), a small population of circulating activated Tfh cells expressing high levels of ICOS, PD-1 and CD38 has been described in peripheral blood of vaccinated individuals for a short period of time post vaccination (Bentebibel et al., 2013; Heit et al., 2017; Herati et al., 2014). In parallel to our FNA analysis, we evaluated activated Tfh cell frequencies in blood of the same immunocompetent subjects vaccinated with SARS-CoV-2 mRNA vaccines. The percentages of blood activated Tfh cells (CD4^+^CD45RA^-^CXCR5^+^ICOS^hi^PD-1^hi^), a large fraction of which also expressed CD38, were significantly increased by SARS-CoV-2 mRNA vaccination (Figure 2H and S2D) and did not correlate with bona fide Tfh cells (gated with a similar strategy) in vaccine draining lymph nodes (Figure 2I). It is worth noting that blood Tfh cells did not correlate with the frequency of SARS-CoV-2-specific GC B cells (Figure S2E), indicating that, although they might reflect the presence of an ongoing GC reaction, they are not accurate biomarkers to estimate bona-fide GC B and Tfh cell responses. Hence GC responses are best studied by direct investigation of vaccine draining lymph nodes.

**Figure 2.**
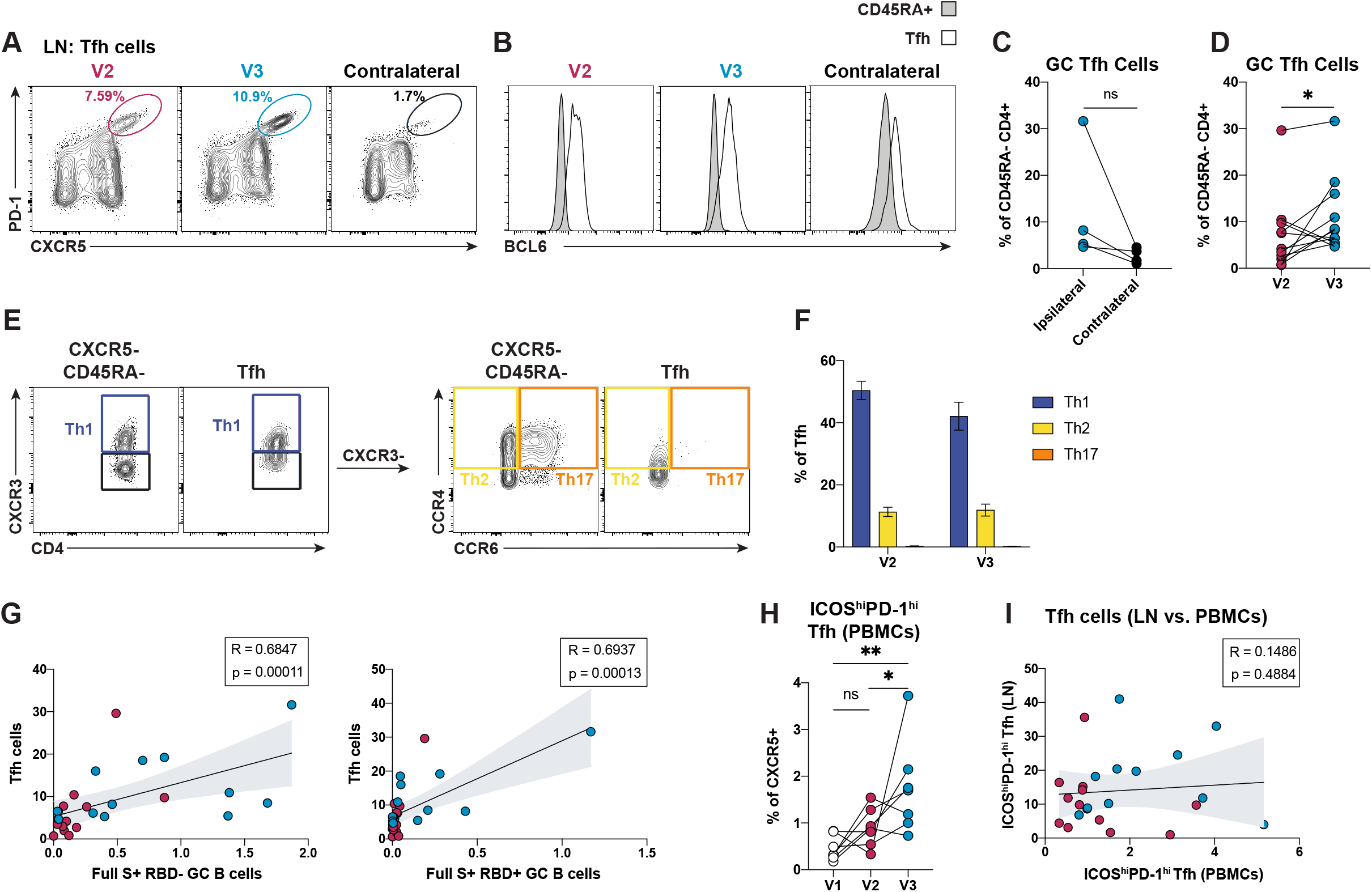
Tfh cell responses with a mixed Th1/Th2 profile are measurable in healthy subject lymph nodes following immunization with SARS-CoV-2 mRNA vaccines. **(A)** Representative flow cytometry plots of Tfh cells (CD4^+^CD8^-^CD19^-^CD45RA^-^CXCR5^hi^PD-1^hi^) in HD FNAs at V2, V3 and from a contralateral lymph node (negative control). **(B)** Expression of BCL6 in Tfh cells from the donors indicated in (A) is displayed as a histogram. **(C)** Quantification of Tfh cells in ipsilateral and contralateral lymph nodes from HDs at V3, displayed as percentage of CD45RA^-^ CD4 T cells. **(D)** Quantification of Tfh cells in ipsilateral lymph nodes from HDs, displayed as a percentage of CD45RA^-^ CD4 T cells. Paired (longitudinal) changes between V2 and V3 are depicted. **(E)** Representative flow cytometry plots for defining Tfh cell subsets. CXCR5^-^CD45RA^-^ (non-Tfh) were used as a control population to set the chemokine receptor gates. **(F)** Quantification of Tfh cell subsets, displayed as a percentage of total Tfh cells. Tfh cells were stratified into: Th1 (CXCR3^+^), Th2 (CXCR3^-^CCR4^+^CCR6^-^), and Th17 (CXCR3^-^CCR4^+^CCR6^+^). Analysis was performed on samples from ipsilateral lymph nodes of HDs at V2 and V3. **(G)** Spearman correlations between Tfh cells (displayed as a percentage of CD45RA^-^ cells) and SARS-CoV-2-specific GC B cells (displayed as a percentage of lymphocytes) from the ipsilateral lymph nodes of HDs at V2 and V3. **(H)** Quantification of activated ICOS^hi^PD-1^hi^ Tfh cells in PBMCs, displayed as a percentage of CXCR5^+^ CD4 T cells. Paired (longitudinal) changes between V1, V2 and V3 are displayed. **(I)** Spearman correlation between activated ICOS^hi^PD-1^hi^ Tfh in PBMCs and bona fide ICOS^hi^PD-1^hi^ Tfh cells from draining lymph nodes (LN) of HDs at V2 and V3, both displayed as a percentage of CXCR5^+^ CD4 T cells. In (C), n = 4; blue data points = ipsilateral lymph node and black data points = contralateral lymph node, both at V3. In (D), n = 11. In (F and G), n = 13 for V2 and V3. In (H), n = 7; white data points = V1, red data points = V2 and blue data points = V3. In (I), n = 13 for V2 and n=11 for V3. In (D, G, and I), red data points = V2 and blue data points = V3. Statistical analysis: In (D and F), a paired (D) and an unpaired (F) Mann-Whitney U test was performed. In (C and H), the Wald-Wolfowitz runs test was performed. In (G and I), correlations were determined using the Spearman’s *rho* with a 95% confidence interval. * P ≤ 0.05, ** P ≤ 0.01.

### Vaccination with SARS-CoV-2 mRNA vaccines leads to the generation of memory B cells and plasmablasts in draining lymph nodes of healthy subjects

As GCs are important for the generation of memory B cells, we next evaluated the memory B cell responses elicited by the two doses of SARS-CoV-2 mRNA vaccines in immunocompetent individuals. Class-switched memory B cells were first defined as IgD^-^IgM^-^CD38^-^CD27^+^ B cells (Figures 3A and S1A). Next, SARS-CoV-2 Full S-specific memory B cells were stratified into RBD^-^ and RBD^+^ cells (Figure 3B), as previously described for GC B cells (Figure 1E). A paired (longitudinal) analysis of matched FNA samples highlighted the generation of Full S and RBD-specific memory B cells after the first vaccine dose, which was significantly higher after the administration of a second mRNA vaccine dose (Figure 3C). An orthogonal analysis approach confirmed similar trends (Figure S3A). SARS-CoV-2-specific memory B cells were also detectable at low frequencies in healthy vaccinee peripheral blood samples after two immunizations (Figure S3B), and only S-specific RBD^-^, but not RBD-specific memory B cell in peripheral blood correlated with the respective SARS-CoV-2-specific memory B cell populations in FNAs (Figure S3C). A population of plasmablasts was also measurable by flow cytometry as IgD^-^IgM^-^CD38^hi^CD20^lo/-^ cells (Figure 3D). This population was more abundant after the booster immunization (Figures 3D-E) and was also detectable at variable levels in peripheral blood samples of vaccinated healthy donors (Figure S3D). However, no correlation was found between the plasmablast populations detected in FNA and blood samples (Figure S3E). In sum, the data obtained in our study indicate that two doses of SARS-CoV-2 mRNA vaccines can elicit SARS-CoV-2 S-and RBD-specific memory B cells as well as a population of plasma cells in vaccine draining lymph nodes.

**Figure 3.**
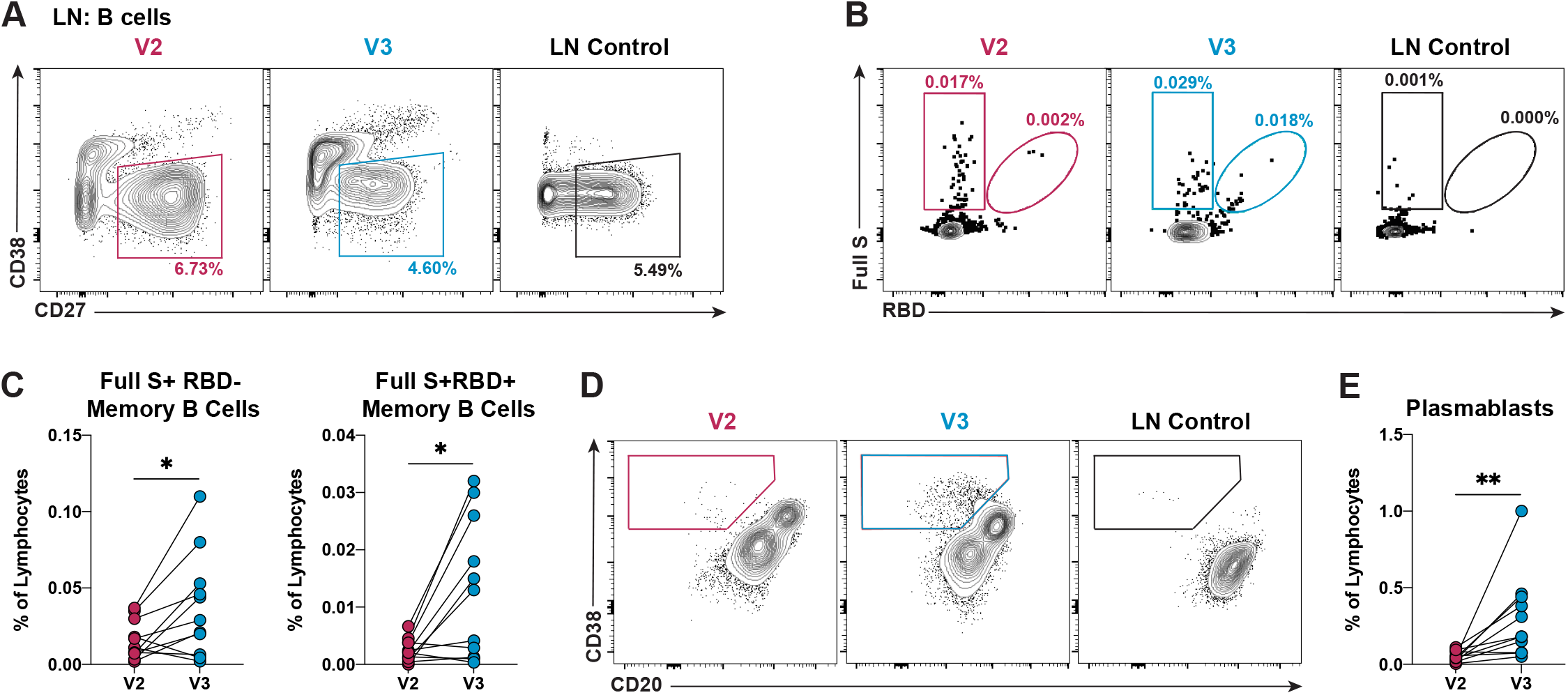
SARS-CoV-2 mRNA vaccinations elicit antigen-specific memory B cell responses. **(A)** Representative flow cytometry of memory B cells (CD19^+^CD4^-^CD8^-^IgM^-^IgD^-^CD38^-^CD27^+^) from ipsilateral lymph nodes of HDs at V2 and V3, or from a putative quiescent cadaveric lymph node (LN Control). **(B)** Representative flow cytometry of SARS-CoV-2-specific memory B cells (HA^-^Full S^+^RBD^-^ or HA^-^Full S^+^RBD^+^) from ipsilateral lymph nodes of HDs at V2 and V3, or a LN control. **(C)** Quantification of Full S^+^ RBD^-^ (**left**) and Full S^+^ RBD^+^ (**right**) memory B cells from ipsilateral lymph nodes of HDs, displayed as a percentage of total lymphocytes. Paired (longitudinal) changes between V2 and V3 are shown. **(D)** Representative flow cytometry of plasmablasts (CD19^+^CD4^-^CD8^-^IgM^-^IgD^-^CD38^hi^CD20^-/lo^) from ipsilateral lymph nodes of HDs at V2 and V3, or a LN control. **(E)** Quantification of plasmablasts in ipsilateral lymph nodes from HDs, displayed as a percentage of total lymphocytes. In (C and E), n = 11; red data points = V2 and blue data points = V3. Statistical analysis: In (C and E), a paired Mann-Whitney U test with continuity correction was performed. * P ≤ 0.05, ** P ≤ 0.01.

### A failure to induce GC B cells by SARS-CoV-2 mRNA vaccines is associated with hindered memory B cell and nAb responses in kidney transplant recipients

We then sought to determine the capacity of immunocompromised individuals to form GC responses to SARS-CoV-2 mRNA vaccines. To this end, 13 individuals who underwent kidney transplant were enrolled. Due to withdrawal of consent (n=1), failure to undergo collection procedures (n=1), and lack of sufficient cells for analysis (n=1), 10 patients who were a median 12.6 months post-transplant (range −0.3-63 months) were included in further analysis (Table 1). Although the draining lymph nodes of KTX recipients were not significantly smaller than HDs (data not shown), the FNA cell recovery yield was scarcer in KTX recipients and, due to limited cell recovery, the immunophenotyping was feasible in this group only at certain time points (Figure S4A). The class-switched B cell populations captured by our 23-color flow cytometry analysis were visualized by a dimensionality-reduction approach (Figure 4A). The most striking observation emerging from this analysis was that a large cell population, reminiscent of GC B cells (CD38^+^CD27^lo/int^BCL6^+^) and present in immunocompetent individuals, was completely lacking in KTX recipients. We further corroborated this finding by a direct evaluation of GC B cell frequencies in KTX patients. This analysis demonstrated an almost complete failure of KTX patients to form GC B cell responses after one or two immunizations with SARS-CoV-2 mRNA vaccines (Figure 4B-C). Importantly, the generation of SARS-CoV-2-specific GC B cells in response to vaccination was completely abrogated even in the few KTX recipients who could mount low but detectable GC B cell responses (Figure 4D).

**Figure 4.**
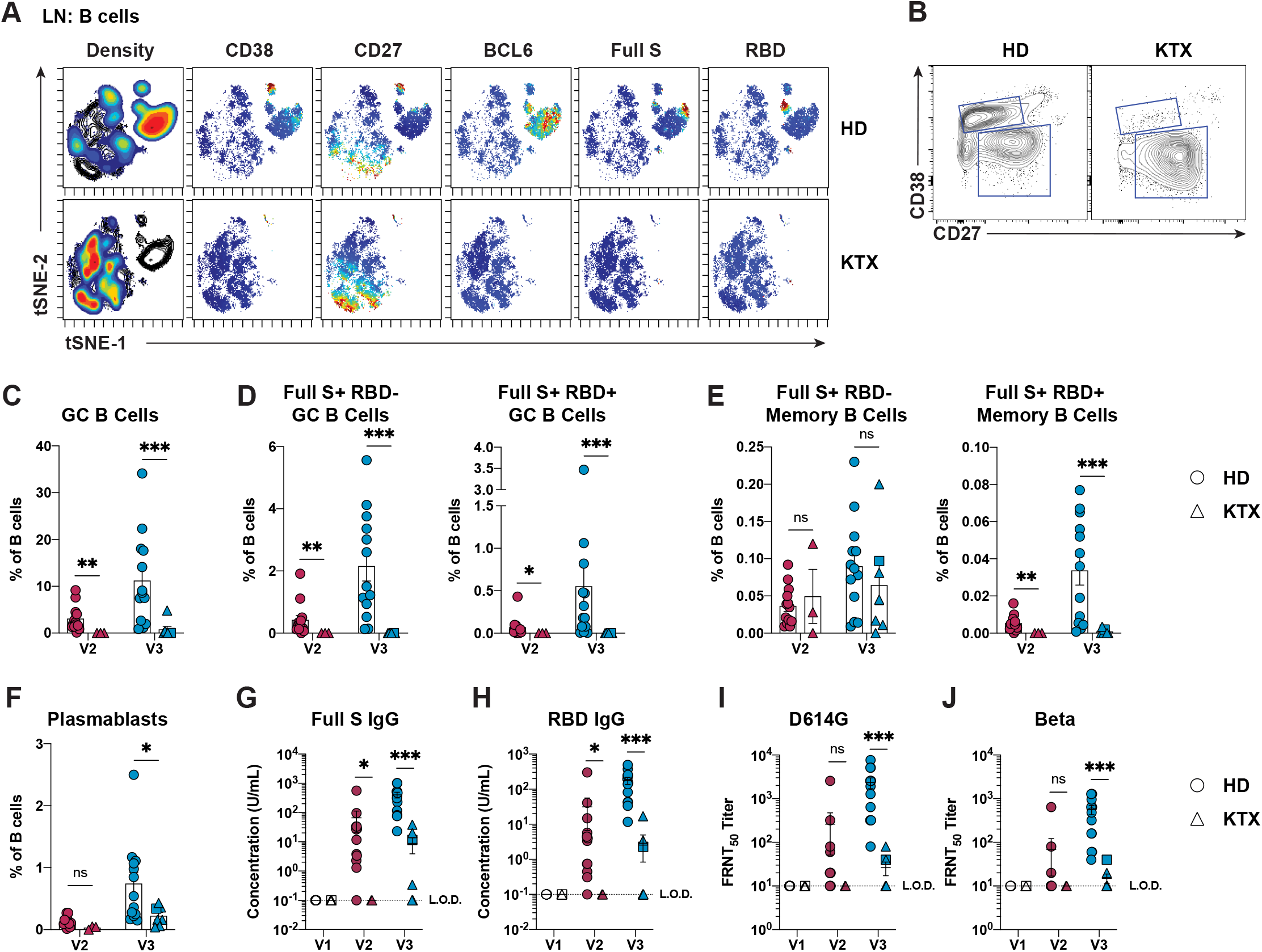
Kidney transplant recipients fail to mount GC reactions and have reduced B cell and humoral responses. **(A)** viSNE analysis of class-switched B cells (CD19^+^CD4^-^CD8^-^IgM^-^IgD^-^) in ipsilateral lymph node samples from HDs and KTX recipients at V3. **(B)** Representative flow cytometry of GC B cells (CD19^+^CD4^-^CD8^-^IgM^-^IgD^-^ CD38^+^CD27^lo/int^BCL6^+^) in ipsilateral lymph node samples from HDs and KTX recipients at V3. **(C and D)** Quantification of GC B cells (C) and SARS-CoV-2-specific GC B cells (D), in ipsilateral lymph node samples from HDs and KTX recipients, displayed as a percentage of B cells. Unpaired (orthogonal) data from V2 and V3 are displayed. **(E)** Quantification of SARS-CoV-2-specific memory B cells, in ipsilateral lymph node samples from HDs and KTX recipients, displayed as a percentage of B cells. Unpaired (orthogonal) data from V2 and V3 are displayed. **(F)** Quantification of plasmablasts, in ipsilateral lymph node samples from HDs and KTX recipients, displayed as a percentage of B cells. Unpaired (orthogonal) data from V2 and V3 are displayed. **(G and H)** Serum concentration of Full S-specific (G) and RBD-specific (H) IgG from HDs and KTX recipients measured by ELISA. **(I and J)** Levels of nAbs against SARS-CoV-2 D614G (I) and Beta (J) mutants measured by pseudoneutralization assay in serum samples from HDs and KTX recipients. In (A), for HD and KTX n=3 at V3. In (C-F), for HD: n = 13 for at V2 and V3; For KTX: n = 3 at V2 and n = 7 at V3. In (G-J), for HD: n = 12 at V1 and V2 and n = 13 at V3; For KTX: n = 7 at V1, n = 2 at V2, and n = 8 at V3. In (C-J), a circle is used to represent HDs, a triangle is used to represent KTX recipients, and a square is used to indicate a KTX recipient with a prior SARS-CoV-2 infection. In (C-J), red data points = V2 and blue data points = V3 and in (G-J) white data points = V1. Statistical analysis: In (C-J), the Wald-Wolfowitz runs test was used to perform an exact comparison between the data distributions for HD versus KTX at each time point. * P ≤ 0.05, ** P ≤ 0.01, *** P ≤ 0.001.

We next questioned whether the dramatic reduction in SARS-CoV-2-specific GC B cells was associated with impaired memory B cell responses to the vaccine, as GCs are an important source for memory B cell production (Mesin et al., 2016). SARS-CoV-2-specific memory B cell frequencies were determined in draining lymph nodes and blood samples. Unexpectedly, the analysis of FNA samples and blood peripheral mononuclear cells (PBMCs) from KTX patients revealed a detectable frequency of Full S^+^RBD^-^ memory B cells within total B cells after the booster immunization as opposed to a complete lack of Full S^+^RBD^+^ memory B cells (Figure 4E and S4B). When analyzed as frequency of memory B cells, however, Full S^+^RBD^-^ memory B cells were decreased in comparison to HDs (Figure S4C). These data, along with the fact that most patients are lymphopenic, indicate that KTX recipients can respond to SARS-CoV-2 mRNA vaccine by producing detectable yet reduced frequencies of Full S-specific memory B cells targeting regions outside RBD. Conversely, they cannot produce measurable RBD-specific memory B cell responses. Interestingly, when HDs and KTX recipients were analyzed together, a strong correlation was found between lymphoid tissue RBD-specific memory B cells and GC B cells, while Full S^+^RBD^-^ memory B cells and GC B cells only presented a weak correlation (Figure S4D). Overall, these intriguing observations suggest that RBD-specific, but not all Full S-specific memory B cells might have a GC origin.

Next, we asked whether and how the absence of vaccine-induced GC B cell responses in KTX recipients might be connected to altered humoral responses, which were previously reported by other groups in a fraction of KTX recipients (Benotmane et al., 2021; Boyarsky et al., 2021a; Kamar et al., 2021; Massa et al., 2021; Stumpf et al., 2021). As a first step in this direction, we evaluated plasmablast frequencies after SARS-CoV-2 vaccine administration. Plasmablast abundance among FNA and PBMC samples was increased after two immunizations with SARS-CoV-2 mRNA in HDs, whereas KTX plasmablast frequencies were, for the most part, reduced in both locations and time points analyzed (Figure 4F and S4E). In line with these data, only ∼40-50% of the KTX recipients in our FNA cohort produced Full S-and RBD-specific IgG within the lower range of HDs after two immunizations, whereas the remaining ∼60-50% of the patients had SARS-CoV-2 binding Abs below the limit of detection (Figure 4G-H). To shed light on the quality of the Ab responses driven by SARS-CoV-2 vaccination in KTX patients after two vaccine doses, we measured SARS-CoV-2 nAbs by pseudotyped lentivirus-based *in vitro* assays. In these assays, the large majority of HD samples collected after two vaccine doses could efficiently neutralize a pseudovirus containing the D614G mutation (Figure 4I) and, less efficiently, a pseudovirus containing the mutation of the SARS-CoV-2 beta strain (Figure 4J). By contrast, KTX patients presented greatly diminished nAbs against D614G-pseudovirus, and KTX plasma could not efficiently block the pseudovirus containing the SARS-CoV-2 beta strain mutations. Correlative analysis including all donors showed that SARS-CoV-2 binding Abs and nAb titers in response to SARS-CoV-2 vaccination were strongly associated with the frequency of Full S RBD^-^ and RBD^+^ GC B cells (Figure S4F-G). Furthermore, while bona fide Tfh cells also displayed a positive correlation with nAb levels, blood activated Tfh cells did not (Figure S4H). Overall, our data demonstrate that KTX recipients cannot mount SARS-CoV-2 specific GC B cell responses or generate RBD-specific memory B cells and efficient nAbs after administration of mRNA vaccines. Furthermore, our work points to a possible connection between GC formation, humoral responses and RBD-specific memory B cell generation in SARS-CoV-2 vaccination.

### Kidney transplant recipients fail to efficiently produce T follicular helper cells and SARS-CoV-2-specific T cell responses

Next, we aimed at determining whether KTX recipients are capable of generating T cell responses to the vaccines, which could counterbalance the impaired B cell responses observed in these patients (Figure 4). As GC B cell responses were heavily impaired in KTX recipients, we predicted reduced Tfh cell frequencies in the KTX group. As anticipated, a viSNE analysis of antigen-experienced CXCR5^+^ CD4 T cells in FNA samples revealed a deep reduction of a cell population expressing the Tfh cell signature markers PD-1, BCL6 and ICOS in the KTX group (Figure 5A). Concordantly, a significant reduction of Tfh cells in KTX patients in comparison to HDs also emerged by a direct flow cytometry analysis (Figures 5B-C).

**Figure 5.**
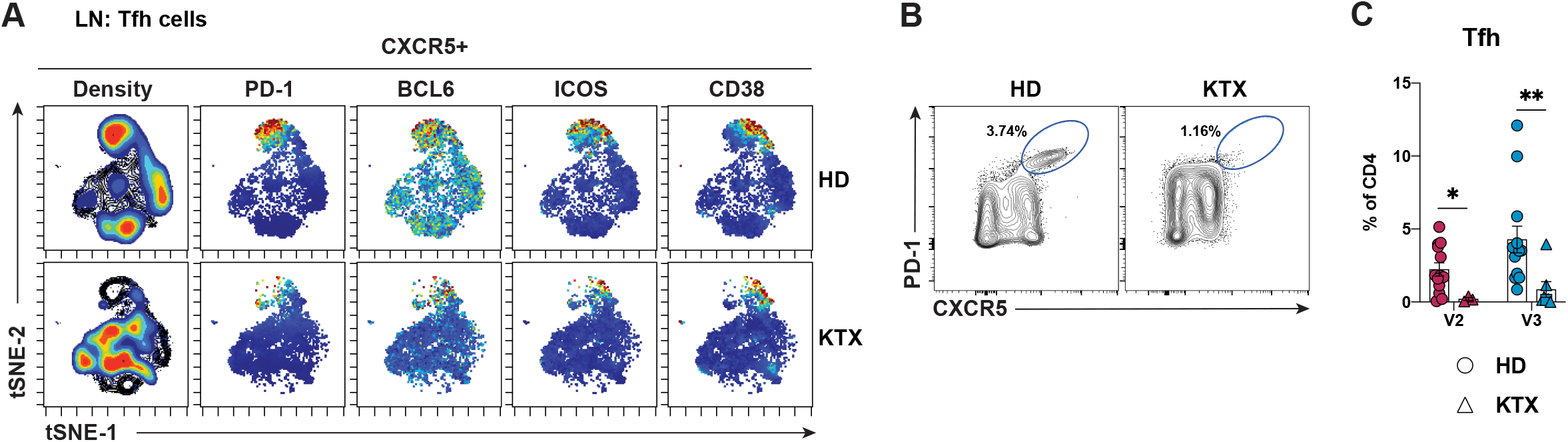
Tfh cell responses to mRNA vaccination are dramatically dampened in kidney transplant recipients. **(A)** viSNE analysis of antigen-experienced CXCR5^+^ CD4 T cells (CD19^-^CD8^-^CD4^+^CD45RA^-^ CXCR5^+^) in ipsilateral lymph node samples from HDs and KTX recipients at V3. **(B)** Representative flow cytometry of Tfh cells (CD4^+^CD8^-^CD19^-^CD45RA^-^CXCR5^hi^PD-1^hi^) from HDs and KTX recipients at V3. **(C)** Quantification of Tfh cells in ipsilateral lymph nodes samples from HDs and KTX recipients shown as a percentage of CD4 T cells. Unpaired (orthogonal) changes between V2 and V3 are displayed. In (A), for HD and KTX n=3 at V3. In (C), for HD: n = 13 for V2 and V3; for KTX: n = 3 for V2 and n = 7 for V3; a circle is used to represent HDs, a triangle is used to represent KTX recipients, and a square is used to indicate a KTX recipient with a prior SARS-CoV-2 infection; red data points = V2 and blue data points = V3. Statistical analysis: In (C), a paired Mann-Whitney U test with continuity correction was performed. * P ≤ 0.05, ** P ≤ 0.01.

We then asked whether KTX recipients are, more broadly, incapable of mounting efficient antigen-specific T cell responses to the SARS-CoV-2 mRNA vaccines. Since a direct evaluation of SARS-CoV-2-specific T cells in vaccine-draining lymph nodes was not feasible due to the paucity and variability in cell recovery of FNAs, we measured the frequency of SARS-CoV-2-specific CD4 and CD8 T cells in peripheral blood of 11 HDs and 10 KTX recipients via an Activation Induced Marker (AIM) assay following *in vitro* stimulation with a SARS-CoV-2 peptide megapool of 253 overlapping 15-mers spanning the Spike protein (Grifoni et al., 2020a). The clinical features of the subjects included in this study are presented in Table 2. Similar to what has previously been reported (Apostolidis et al., 2021; Painter et al., 2021), we detected significantly increased frequencies of AIM^+^ (CD200^+^CD40L^+^) SARS-CoV-2-specific CD4 T cells in blood samples of all HDs after the first vaccine administration (Figure 6A-B and Figure S5A). These appeared to be further boosted by the second vaccine administration, as has been observed in other studies (Apostolidis et al., 2021; Painter et al., 2021), although this did not reach statistical significance in our cohort. In contrast, we observed a severely reduced induction of antigen-specific CD4 T cells in KTX recipients after either vaccine administration. Since KTX recipients might present altered ratios of naïve/antigen-experienced CD4 T cells, we also analyzed the AIM^+^ CD4 T cells as frequency of total CD4 T cells, observing a similar attenuation of SARS-CoV-2-specific CD4 T cells in the KTX group compared to HDs (Figure S5B). A functional stratification of the AIM^+^ CD4 T cells based on chemokine receptor expression allowed us to identify SARS-CoV-2-specific circulating Tfh cells (CXCR5^+^), as well as Th1 (CXCR3^+^), Th17 (CCR6^+^) and Th2 (CXCR3^-^ CCR6^-^)-polarized CXCR5^-^ non-Tfh cells (Figure 6C) (Acosta-Rodriguez et al., 2007; Morita et al., 2011; Trifari et al., 2009). As previously observed, healthy individuals predominantly generate SARS-CoV-2-specific circulating Tfh and Th1 polarized CD4 T cells in response to the mRNA vaccines (Figure 6C-D) (Apostolidis et al., 2021; Painter et al., 2021). Low frequencies of antigen-specific Th2-biased CD4 T cells were also present. Of note, the SARS-CoV-2-specific circulating Tfh cells detected in HDs via the AIM assay presented a mixed Th1/Th2 functional polarization (Figure 6E) similar to what was observed in draining lymph node bona fide Tfh cells (Figure 2F).

**Figure 6.**
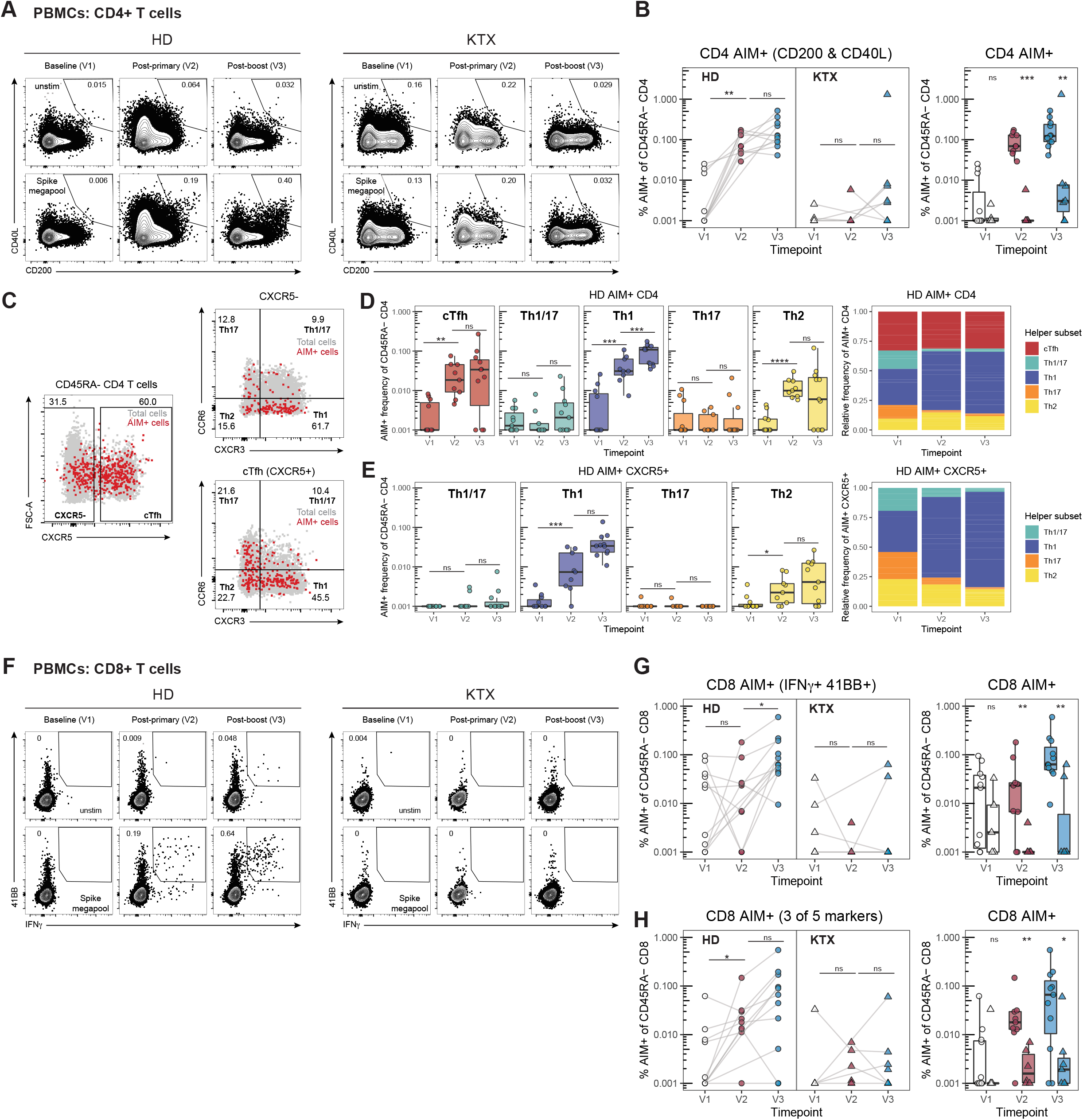
Kidney transplant recipients fail to generate effective antigen-specific T cell responses. **(A)** Representative flow cytometry of AIM^+^ (CD200^+^CD40L^+^) CD4 T cells in PBMCs at V1, V2 and V3 from HDs and KTX recipients. **(B)** Quantification of AIM^+^ CD4 T cells, defined as in (A), displayed as a percentage of antigen-experienced (CD45RA^-^) CD4 T cells. Paired (longitudinal, left) or unpaired (orthogonal, right) analyses of PBMC samples from HDs and KTX recipients at V1, V2 and V3 are shown. **(C)** Representative gating strategy to define AIM^+^ CD4 T cell subsets in HD PBMC samples. **(D and E)** Quantification of AIM^+^ total CD4 T cell subsets (D) and AIM^+^ CXCR5^+^ CD4 T cell subsets (E) in HDs. Unpaired (orthogonal) analysis of PBMC samples from V1, V2 and V3 in HDs is displayed. **(F)** Representative flow cytometry of AIM^+^ (IFNγ^+^ and 41BB^+^) CD8 T cells in PBMCs at V1, V2 and V3 from HDs and KTX recipients. **(G)** Quantification of AIM^+^ (IFNγ^+^ and 41BB^+^) CD8 T cells, displayed as a percentage of antigen-experienced (CD45RA^-^) CD8 T cells. Paired (longitudinal, left) or unpaired (orthogonal, right) analyses of PBMC samples from HDs and KTX recipients at V1, V2 and V3 are shown **(H)** Quantification of AIM^+^ (cells expressing at least 3 of 5 activation markers: CD107a, 41BB, CD200, CD40L, and IFNγ) CD8 T cells, displayed as a percentage of antigen-experienced (CD45RA^-^) CD8 T cells. Paired (longitudinal, left) or unpaired (orthogonal, right) analyses of PBMC samples from HDs and KTX V1, V2 and V3 recipients are shown. In (B, D-E and G-H), for HD: n = 11 for V1and V3, n = 9 for V2; for KTX: n = 5 for V1, n = 6 for V2, and n = 7 for V3. In (B and G-H), a circle is used to represent HD and a triangle is used to represent KTX; white data points = V1, red data points = V2 and blue data points = V3. Statistical analysis: In (B-H), the Wald-Wolfowitz runs test was used to perform an exact comparison between the two data distributions of interest. * P ≤ 0.05, ** P ≤ 0.01, *** P ≤ 0.001, **** P ≤ 0.0001.

In the few KTX recipients that had detectable AIM^+^ cells after two vaccinations, we did not observe major alterations in the functional polarization of SARS-CoV-2-specific CD4 T cells (Figure S5C-D), though the small numbers of AIM^+^ CD4 T cells in these donors prevented us from making definitive conclusions. The analysis of SARS-CoV-2-specific CD8 T cells followed a similar trend to that observed for CD4 T cell responses. SARS-CoV-2-specific CD8 T cells, defined either as CD8 T cells co-expressing 41BB and IFNγ or expressing 3 out of 5 AIM markers used in our panel, were variable in frequency but detectable in most healthy vaccinees above pre-vaccine baseline levels (Figures 6F-H and S5E-F). In contrast, most KTX recipients did not present detectable SARS-CoV-2-specific CD8 T cells, resulting in significantly reduced responses compared to HDs at both post-vaccine time points (Figure 6F-H and S5E-F). As opposed to the altered antigen-specific T cell responses, we observed similar frequencies of total (non-antigen-specific) CD4 T cell subsets across all time points in both HDs and KTX recipients (Figure S5G-H). Altogether, these data point to severely decreased SARS-CoV-2-specific CD4 and CD8 T cell responses in KTX after two immunizations with SARS-CoV-2 mRNA vaccines.

## DISCUSSION

mRNA vaccines are a novel vaccine platform that has only been recently approved for human use during the current coronavirus disease 2019 (COVID-19) pandemic. While it is emerging that SARS-CoV-2 mRNA vaccines are highly efficient at inducing robust nAbs and memory B cell responses, we still have limited knowledge of the underlying mechanisms leading to the generation of such immune responses in humans. Specifically, a fundamental open question is whether nAb and memory B cell generation during SARS-CoV-2 mRNA vaccination is connected to the formation of GCs, microanatomical structures in secondary lymphoid organs harboring the generation of affinity matured Ab-secreting cells and memory B cells. The study of vaccine-induced GC reactions in humans is heavily constrained when blood, the most easily obtainable human material, is the only available sample because bona fide GC B cells and Tfh cells are only present in secondary lymphoid organs (Vella et al., 2019). Surrogate biomarkers such as blood CXCL13 and circulating activated Tfh cells have been used thus far to predict the magnitude of ongoing GC responses (Havenar-Daughton et al., 2016; Vella et al., 2019). However, these biomarkers present significant shortcomings when trying to directly assay GC responses including the fact that they are traceable for only short windows of time in blood, are not detectable in all individuals in response to vaccination, and have not been successfully used to fully predict broad qualitative aspects of GC reactions, such as the antigen specificity of GC B cells and their connection to GC-derived B cell responses (Bentebibel et al., 2013; Havenar-Daughton et al., 2016). These limitations emphasize the need for directly probing GC responses by adopting minimally invasive approaches, such as fine-needle-aspiration, that allow longitudinal sampling of the vaccine draining lymph nodes without requiring their surgical excision. Only two published studies ever described human GC responses to vaccination (against influenza or SARS-CoV-2) in humans by using the FNA technique (Turner et al., 2020, 2021). One study (Turner et al., 2021), reported S-specific GC B cell responses to SARS-CoV-2 mRNA vaccination in lymph nodes, but did not investigate the generation of RBD-specific GC B cells or the connection between SARS-CoV-2-specific GC B cells and Tfh cells, nAbs, and SARS-CoV-2-specific memory B cells. Our study sought to address these open questions by performing an in-depth profiling of the GC responses elicited by SARS-CoV-2 mRNA vaccines directly in lymphoid tissue. We found that these vaccines prompted the formation of robust Full S and RBD-specific GC B cell as well as Tfh cell responses localized in draining axillary lymph nodes. Importantly, our study revealed that lymphoid tissue SARS-CoV-2-specific GC B cell populations were strongly associated with the ability to produce SARS-CoV-2 nAbs, as further supported by the evidence that immunosuppressed individuals, who cannot form GCs, present a deeply blunted nAb production (Results in KTX are discussed at length below). Although to a lower degree, we also found that bona fide Tfh cells in draining lymph nodes correlated with nAb production, while activated Tfh cells in blood were a less reliable predictor of nAb generation. Additionally, in our study GC formation appeared to be tightly connected with the capacity to produce RBD-specific memory B cells. These findings, which we would have been unable to observe by studying blood alone, provide valuable insights on the otherwise poorly understood processes by which nAbs and memory B cells are formed in humans after immunization with SARS-CoV-2 mRNA vaccines.

SARS-CoV-2-specific Abs, including nAbs, play a key role in the protection against COVID-19, as indicated by passive transfer experiments in animal models (McMahan et al., 2021; Rogers et al., 2020; Zost et al., 2020) and prospective cohort studies in previously-infected or vaccinated humans (Bergwerk et al., 2021; Earle et al., 2021; Khoury et al., 2021; Lumley et al., 2020). Herein, we have observed a strong association between SARS-CoV-2-specific GC B cells and nAbs, which suggests that GC responses are critical to mount a SARS-CoV-2 nAb response during vaccination. In line with this connection, we have previously shown that animals immunized with SARS-CoV-2 mRNA vaccines efficiently formed RBD-specific GC B cells coupled with robust nAbs levels, as opposed to mice immunized with a protein RBD antigen formulated in an MF59-like adjuvant, which mounted negligible GC responses and subsequently formed limited nAbs (Lederer et al., 2020). It is worth noting that a similar connection between GC formation and nAb production has not been observed during SARS-CoV-2 infection. Multiple groups have reported that several near-germline nAbs, endowed with potent *in vitro* neutralizing activity, are elicited during natural SARS-CoV-2 infection in humans (Brouwer et al., 2020; Kreer et al., 2020; Schultheiß et al., 2020; Seydoux et al., 2020). The low degree of SHM of these Abs is suggestive of a limited GC process involved in their generation. Similarly, a pre-print from Eisenbarth and colleagues shows that Tfh cell-deficient mice, which form negligible GCs in response to SARS-CoV-2 infection, present decreased yet detectable production of Abs that can neutralize SARS-CoV-2 *in vitro* (Chen et al., 2021). Overall, these data indicate that GCs might not be necessary to form nAbs in response to infection. This apparently discordant outcome might stem from a combination of diverse factors, including a different pool of germline B cells recruited by SARS-CoV-2 mRNA vaccination, but not during natural infection. Both BNT162b2 or mRNA-1273 vaccines encode for a prefusion stabilized version of SARS-CoV-2 Full S protein (Wrapp et al., 2020) that induce nAbs more targeted to the RBD and that bind more broadly across the RBD in comparison to the infection-induced nAbs (Greaney et al., 2021). The broader binding suggests that, in comparison to natural infection, additional/alternative germline precursors of the nAb secreting cells are recruited by SARS-CoV-2 mRNA vaccines. It is tempting to speculate that some of these nAb precursors require GC reactions to produce high-affinity nAbs, as supported by the finding that plasmablasts emerging from SARS-CoV-2 mRNA vaccination can present high levels of SHM (Amanat et al., 2021). As alternative or complementary explanation for the GC-nAb connection described in our current and previous studies, the mRNA vaccine platform might favor the formation of GC-derived nAbs thanks to its strong pro-GC activity. We and others have shown that the mRNA vaccine platform is very effective at promoting the formation of GC reactions in animal models (Lederer et al., 2020; Lindgren et al., 2017; Pardi et al., 2018a), with a mechanism that relies on an early induction of the pro-Tfh cytokine IL-6 by the lipid nanoparticle component of these vaccines (manuscript under revision). The present study, along with the recently published work by Ellebedy and colleagues (Turner et al., 2021), further extend our earlier observation (Lederer et al., 2020) by directly showing the formation of SARS-CoV-2 specific GC B cells in humans after mRNA vaccination. Hence, by eliciting effective GC responses and/or potentially recruiting nAb precursors that can seed GCs, SARS-CoV-2 mRNA vaccines might mechanistically rely on GC responses to effectively generate nAbs, as strongly supported by the associations between GCs and nAbs found in this study.

The second part of our study was aimed at evaluating the GC responses in KTX recipients and their association with humoral and memory B cell responses, as no study has ever evaluated B cell responses in their place of origin (vaccine-draining lymphoid tissue) in immunocompromised individuals. In agreement with other studies reporting that a fraction of KTX recipients can still produce SARS-CoV-2-binding Abs after two immunizations with SARS-CoV-2 mRNA vaccines (Benotmane et al., 2021; Boyarsky et al., 2021a; Kamar et al., 2021; Massa et al., 2021; Stumpf et al., 2021), we observed that 40-50% of the KTX recipients enrolled in our FNA study produced Full S-and RBD-specific IgG, albeit within the lower range of healthy controls. This finding was coupled to the observation that several KTX recipients had low but detectable plasmablast frequencies following the booster immunization. Of note, we also found that KTX recipients can generate reduced but detectable vaccine-induced SARS-CoV-2-specific memory B cells targeting the Full S protein outside the RBD region. Nonetheless, our study reported for the first time a complete failure of KTX recipients in forming lymphoid tissue SARS-CoV-2-specific GC B cell responses after the administration of two mRNA vaccine doses. This finding was accompanied by a markedly diminished generation of RBD-specific memory B cells and SARS-CoV-2 nAbs. Two other groups reported low SARS-CoV-2 vaccine-induced nAb levels in large SOT cohorts (Massa et al., 2021; Rincon-Arevalo et al., 2021) and decreased presence of class-switched RBD^+^ B cells (Rincon-Arevalo et al., 2021) in peripheral blood induced by two vaccine doses. Our study confirms and extends this observation by suggesting that defective GC formation could be the underlying culprit behind the decreased nAb and lack of RBD-specific memory B cell production in KTX recipients. Overall, our study indicates that, while some residual immune response is obtainable by SARS-CoV-2 mRNA vaccination, B cell responses appear to be quantitatively and qualitatively curtailed in these immunocompromised subjects.

Given the suboptimal humoral and B cells responses of KTX recipients following SARS-CoV-2 mRNA vaccination, an important question is whether mRNA vaccines are capable of eliciting T cell responses in immunocompromised individuals. In immunocompetent subjects, the licensed mRNA vaccines predominantly promote the formation of circulating blood Tfh cells and Th1-polarized CD4 T cells (Painter et al., 2021). While a limitation of our study was the inability to assess SARS-CoV-2-specific T cell responses in vaccine draining lymph nodes due to the scarcity of available material, we observed that KTX recipients were almost completely deprived of bona fide lymph node Tfh cells after SARS-CoV-2 mRNA vaccination, which is consistent with the lack of GC B cell formation in these patients that is also described in this study. In parallel, we observed, consistent with other studies (Sattler et al., 2021; Stumpf et al., 2021), that SARS-CoV-2-specific CD4 (including circulating Tfh cells and Th1-polarized cells) and CD8 T cell populations were decreased in frequency in KTX recipients when compared to the HD group. Overall, our study indicates that KTX recipients poorly respond to two immunizations with SARS-CoV-2 mRNA vaccines by failing to produce efficient humoral and cellular responses, with some immune responses (GC B cells, nAb RBD-specific memory B cells and SARS-CoV-2-specific T cells) that were more severely affected than others (binding Abs, Full S-specific memory B cells). Since an increasing body of evidence is now suggesting that the administration of a third vaccine dose can significantly boost SARS-CoV-2 binding Abs and nAbs in recipients of SOT (Hall et al., 2021; Kamar et al., 2021; Massa et al., 2021), it will be interesting to assess in the future whether this is due to the expansion of the small pool of preexisting (Full S^+^RBD^-^) memory B cells that we identified in this study, or to an increased *de novo* generation of Abs via GC reactions.

Suboptimal vaccine responses in organ transplant subjects receiving immunosuppressant drugs was previously reported for other vaccines including influenza A/H1N1 and Hepatitis B (Brakemeier et al., 2012; Broeders et al., 2011; Cowan et al., 2014; Elhanan et al., 2018; Friedrich et al., 2015). Collectively, these studies reported impaired Ab production post vaccination, which is reflective of a dysfunction of the immune system in individuals receiving immunosuppressant drugs. The heavily diminished induction of GC B cell, memory B cell, nAb, and T cell responses in our KTX patients reported by this study is likely a consequence of lymphopenia as well as immunosuppression-induced immune cell dysfunction. KTX patients in this study uniformly received anti-thymocyte globulin (ATG) as induction immunosuppression at the time of kidney transplantation (completed within a week). Hence, an incomplete reconstitution of the T cell pool following ATG administration might at least partially explain the hampered Tfh and SARS-CoV-2-specific T cell responses in patients in our cohort who were recently transplanted. Additionally, maintenance immunosuppression comprised prednisone, a calcineurin inhibitor and an anti-metabolite. Indeed, the majority of patients were lymphopenic, as evaluated by the baseline absolute lymphocyte count (8/9 in the FNA cohort, 9/11 in the blood cohort), as a consequence of ATG and/or maintenance immunosuppression. Currently, there is discordant data with regard to the impact of individual immunosuppressive drugs on the immune response to SARS-CoV-2 mRNA vaccines in SOT populations (Boyarsky et al., 2021a; Cucchiari et al., 2021; Grupper et al., 2021), likely due to the diversity of pharmacologic/biologic immunosuppressive agents and doses used. Due to the limited size of our cohorts, we were unable to meaningfully attribute impacts of any of the individual agents on the GC process. Future studies with larger cohorts of KTX recipients will be needed to address the relative contribution of ATG and immunosuppressant drugs to the disrupted GC formation that we observed in these patients.

In sum, by directly probing GC responses at their source, we provided a unique perspective on the connection between GC formation and nAb/memory B cell generation following immunizations with SARS-CoV-2 mRNA vaccines in healthy and immunocompromised individuals. Broadly, this work will pave the road to future human vaccine studies aimed at untangling the origin of long-lasting, protective immune responses after immunizations with different licensed-vaccines.

## Data Availability

All data generated or analyzed during this study are included in this published article or available from the corresponding author upon (reasonable) request.

## ACKNOWLEDGEMENTS

M.L. was supported by NIH NIAID grants R01 AI123738 and R01AI153064. This work was funded by The Gift of Life Transplant Foundation (V.B. and A.N.), the National Blood Foundation (V.B.), the Burroughs Welcome Fund (V.B.), the U19AI082630 (S.E.H. and E.J.W.) and by the NIH NIAID under contract Nr. 75N9301900065 (D.W., A.S.). First, the authors wish to thank all of the subjects for their participation in this study. The authors also thank Dr. Fatima Amanat and Dr. Florian Krammer for kindly providing the RBD protein used in this study, Diane Mclaughlin and Sarah Benchimol for administrative assistance, Susan Rostami for assistance with sample processing, Jennifer Trofe-Clark and Gregory Malat for regulatory assistance and Moses Awofolaju, Nicole Tanenbaum, and Jordan Ort for assistance with ELISAs. We thank Dr. Florin Tuluc and Jennifer Murray of the CHOP Flow Cytometry core facility for technical assistance and the Flow Cytometry Core at the University of Pennsylvania.

## AUTHOR CONTRIBUTIONS

K.L., E.B. and M.L. performed and/or analyzed the immunophenotyping of FNA and blood samples. M.P, K.P. and R.G. performed and/or analyzed AIM assays. D.A. performed statistical analysis. K.A.L. and P.B. performed and/or analyzed the neutralization assays. M.W., E.M.D., S.G and S.E.H. performed and/or analyzed serological data. X.X. processed blood samples. A.G. provided support with Cytobank analysis. C.LC. and N.R. provided tonsil samples. L.J. and M.R. shared expertise for FNA procedures. D.W. and A.S. provided SARS-CoV-2 peptide mega-pools. A.N., M.K., B.B., V.B. and K.P. supervised the recruitment of the subjects involved in the study as well as FNA and blood sample collection. M.L. wrote the manuscript with help from K.L, E.B. and V.B. and input from the other authors. M.L conceived and supervised the study with support from E.J.W., A.N. and V.B.

## DECLARATION OF INTERESTS

EJW is consulting or is an advisor for Merck, Elstar, Janssen, Related Sciences, Synthekine and Surface Oncology. EJW is a founder of Surface Oncology and Arsenal Biosciences. EJW is an inventor on a patent (US Patent number 10,370,446) submitted by Emory University that covers the use of PD-1 blockade to treat infections and cancer. S.E.H. has received consultancy fee from Sanofi Pasteur, Lumen, Novavax, and Merck for work unrelated to this report. A.S. is a consultant for Gritstone, Flow Pharma, Arcturus, Immunoscape, CellCarta, Oxford Immunotech and Avalia.

## METHODS

### EXPERIMENTAL MODEL AND SUBJECT DETAILS

#### Study design and human samples

The prospective cohort study included 15 healthy adults and 14 kidney transplant recipients at the Hospital of the University of Pennsylvania across both the lymph node and blood samples analyses (**Table 1 and 2)**. All participants received two doses of either BNT162b2 or mRNA-1273 vaccines, according to the recommended 3-and 4-week interval, respectively. All participants received the first and second immunizations in the same arm. Written informed consent for participation was obtained according to the Declaration of Helsinki and protocols were approved by the Institutional Review Board of the University of Pennsylvania. Lymph node samples were obtained by ultrasound-guided fine needle aspiration at day 12 (+/- 3 days) after primary immunization and at day 10 (+/- 2 days) after booster immunization. Blood samples were obtained at baseline prior to vaccination (visit 1, V1), day 12 (+/- 3 days) following primary immunization (visit 2, V2), and day 10 (+/- 2 days) following booster (visit 3, V3).

Lymph nodes and pediatric tonsils were obtained from the National Disease Resource Interchange (NDRI), and the Children’s Hospital of Philadelphia (CHOP), respectively.

#### Ultrasound guided fine needle aspiration

All fine needle aspirations (FNA) were performed by board-certified radiologists, similar to what previously described (Havenar-Daughton et al., 2020). Briefly, a Philips EPIQ ELITE or PHILIPS IU222 ultrasound instrument was used to visualize axillary draining lymph nodes. The area around the lymph node was anesthetized using 2-6mL of 0.9% buffered lidocaine solution. A 25-gauge needle was inserted into the cortex and moved back-and-forth several times, sample was aspirated and ejected into cold RPMI media containing 10% FBS. A total of five such passes were performed. In all participants, FNAs were performed on the side of vaccination (ipsilateral). In 4 participants, additional FNA was performed on the contralateral side.

#### Blood processing

##### Isolation of serum

Blood was collected in serum separator tubes (Becton Dickinson) which were spun at 935g for 15 minutes. Serum was collected, aliquoted, and frozen at −80°C for subsequent use.

##### Isolation of Peripheral Blood Mononuclear Cells (PBMCs)

PBMCs were isolated from blood collected in sodium heparin vacutainer tubes (Becton Dickinson). Briefly, whole blood was first spun at 935g for 15min. The plasma was carefully collected, aliquoted and stored at −80°C. The buffy layer and red cell sediment were diluted with an equal volume of RPMI with 5% FBS (RPMI-5) and gently layered over 15mL of Ficoll-Paque Plus (Cytiva) in a 50mL SepMate tube (STEMCELL Technologies). The sample was centrifuged at 1200g for 10 minutes. The PBMC were transferred into a new 50mL conical tube, centrifuged, decanted, and washed twice with RPMI-5 before flow cytometry staining or cryopreservation in FBS with 10% dimethyl sulfoxide.

#### Production of fluorescently labeled proteins

##### Labeling of SARS-CoV-2 full-length spike protein

Full-length, biotinylated spike protein was purchased from R&D Systems. Streptavidin-conjugated BV421 (Biolegend) was then added at a 6:1 molar ratio (biotinylated-protein to streptavidin-conjugate) on ice for 1 hour.

##### Labeling of HA and SARS-CoV-2 RBD

Recombinant HA and RBD was produced as previously described (Amanat et al., 2020; Margine et al., 2013; Stadlbauer et al., 2020). To create fluorescently labeled RBD tetramers, RBD was biotinylated using the EZ-Link Micro Sulfo-NHS-Biotinylation Kit (ThermoFisher). Streptavidin-conjugated PE was then added at a 6:1 molar ratio (biotinylated-protein to streptavidin-conjugate). Specifically, after the volume of fluorochrome needed to achieve a 6:1 molar ratio was determined, the total volume of fluorochrome was split into 10 subaliquots. These subaliquots were then added, on ice, to the biotinylated protein and mixed by pipetting every 10 minutes (for a total of 10 additions).

#### Flow cytometry

Staining was performed on freshly isolated FNA and PBMC samples or cryopreserved control LN and tonsil samples. Up to 10^6^ cells were incubated with a cocktail of chemokine receptor antibodies in FACS buffer (PBS containing 2% FBS and 1mM EDTA) for 10 minutes at 37℃. All remaining steps were carried out at 4℃. Without washing, a 2x cocktail of all other surface antibodies diluted in Brilliant Violet Staining Buffer (BD Biosciences) was added directly and incubated for 1 hour. Cells were washed with FACS buffer, fixed and permeabilized with FoxP3 Fixation/Permeabilization Buffer (eBioSciences) according to manufacturer’s instructions for 1 hour, and incubated with anti-BCL6 mAb (BD Biosciences) for 30 minutes. The 23-color panel used in this study is described in Table 3. Samples were washed, resuspended in FACS buffer and immediately acquired on an Aurora using SpectroFlow v2.2 (Cytek). Data was analyzed using Flow v.10 (Treestar).

#### viSNE analysis

viSNE analysis was performed on Cytobank (https://cytobank.org).

##### Class-switched B cell analysis

Cells were defined as live, CD8^-^, CD4^-^, CD19^+^, IgD^-^ IgM^-^. viSNE analysis was performed using 3200 cells from n = 3 donors per cohort with 5000 iterations, a perplexity of 60 and a theta of 0.5. The following markers and/or probes were used to generate viSNE projections: CXCR5, CD11b, CD11c, CD20, CD27, CD38, BCL6, CCR4, CCR6, CXCR3, CD138, ICOS, PD-1, RBD Probe, Full S Probe, HA Probe.

##### CXCR5^+^ CD4 T cell analysis

Cells were defined as live, CD8^-^, CD19^-^, CD4^+^, CD45RA^-^ CXCR5^+^. Analysis was performed using 2730 cells from n = 3 donors per cohort with 5000 iterations, a perplexity of 100 and a theta of 0.5. The following markers and/or probes were used to generate viSNE projections: PD-1, BCL6, CCR4, CCR6, CXCR3, CD11b, CD11c, CD20, CD27, CD38, ICOS.

#### Activation induced marker (AIM) expression assay

The AIM assay was performed as previously described (Painter et al., 2021). Briefly, after thawing and counting, cells were resuspended in fresh R10 to a final density of 10×10^6^ cells/mL, and 2×10^6^ cells in 200μL were plated in duplicate wells in 96-well round-bottom plates. After resting overnight, CD40 blocking antibody was added to both duplicate wells for 15 minutes prior to stimulation. One of the duplicate wells was then stimulated for 24 hours with costimulation (anti-human CD28/CD49d, BD Biosciences) and the Spike peptide megapool at a final concentration of 1 mg/mL, while the other well was treated with costimulation alone as a paired unstimulated sample. The CD4-S peptide megapool consists of 253 overlapping 15-mer peptides spanning the entire sequence of the Spike protein and was prepared as previously described (Grifoni et al., 2020b, 2020a). The remainder of the AIM assay was performed and samples were collected and analyzed as previously described (Painter et al., 2021). The flow cytometry panel used for the the detection of AIM^+^ cell populations is described in Table 4.

AIM^+^ cells were identified from non-naïve or total T cell populations where indicated. All data from AIM expression assays were background-subtracted using paired unstimulated control samples. For T cell subsets, the AIM^+^ background frequency of CD45RA^-^ T cells was subtracted independently for each subset. AIM^+^ CD4 T cells were defined by dual-expression of CD200 and CD40L. AIM^+^ CD8 T cells were defined by either expression of 41BB and IFNγ or a boolean analysis identifying cells expressing at least three of five markers: CD200, CD40L, 41BB, CD107a, and intracellular IFNγ.

AIM assay data were visualized using RStudio. Boxplots represent median with interquartile range. Source code and data files are available upon request from the authors.

#### Enzyme-linked immunosorbent assay

ELISAs were performed using a previously described protocol (Flannery et al., 2020). Plasmids expressing the receptor binding domain (RBD) of the SARS-CoV-2 spike protein and the full-length (FL) spike protein were provided by F. Krammer (Mt. Sinai). SARS-CoV-2 RBD and FL proteins were produced in 293F cells and purified using nickel–nitrilotriacetic acid (Ni-NTA) resin (Qiagen). The supernatant was incubated with Ni-NTA resin at room temperature for 2 hours before collection using gravity flow columns and protein elution. After buffer exchange into phosphate-buffered saline (PBS), the purified protein was aliquoted and stored at −80°C. ELISA plates (Immulon 4 HBX, Thermo Fisher Scientific) were coated with PBS (50 µl per well) or a recombinant SARS-CoV-2 RBD or FL proteins (2 µg/ml) diluted in PBS and stored overnight at 4°C. ELISA plates were washed three times with PBS containing 0.1% Tween 20 (PBS-T) and blocked for 1 hour with PBS-T containing 3% nonfat milk powder. Serum samples that had been previously heat-inactivated (56°C for 1 hour) were serially diluted four-fold in 96-well round-bottom plates in PBS-T supplemented with 1% nonfat milk powder (dilution buffer), starting at a 1:50 dilution. ELISA plates were then washed three times with PBS-T. 50 µl of serum dilution was added to each well and incubated at room temperature for 2 hours. Plates were then washed again with PBS-T three times and 50 µl of horseradish peroxidase (HRP)–labeled goat anti-human IgG (1:5000; Jackson ImmunoResearch Laboratories) secondary antibodies was added. After 1-hour incubation at room temperature, plates were washed three times with PBS-T, 50 µl of SureBlue 3,3′,5,5′-tetramethylbenzidine substrate (KPL) was added to each well, and 25 µl of 250 mM hydrochloric acid was added to each well to stop the reaction five minutes later. Plates were read at an optical density (OD) of 450 nm using the SpectraMax 190 microplate reader (Molecular Devices). All incubation and washing steps were performed using a plate mixer. For analyses, OD values from the plates coated with PBS were subtracted from the OD values from plates coated with either RBD or FL recombinant protein, to control for background ELISA antibody binding. Each plate contained a dilution series of the IgG monoclonal antibody CR3022, which is reactive to the SARS-CoV-2 spike protein, to control for variability between assays. Serum antibody concentrations were reported as arbitrary units relative to the CR3022 monoclonal antibody.

#### Pseudovirus neutralization assay

##### Production of VSV pseudotypes with SARS-CoV-2 S

293T cells plated 24 hours previously at 5 × 10^6^ cells per 10 cm dish were transfected using calcium phosphate with 35 µg of pCG1 SARS-CoV-2 S D614G delta18 or pCG1 SARS-CoV-2 S B.1.351 delta 18 expression plasmid encoding a codon optimized SARS-CoV2 S gene with an 18 residue truncation in the cytoplasmic tail (kindly provided by Stefan Pohlmann). Twelve hours post transfection the cells were fed with fresh media containing 5mM sodium butyrate to increase expression of the transfected DNA. Thirty hours after transfection, the SARS-CoV-2 spike expressing cells were infected for 2-4 hours with VSV-G pseudotyped VSVΔG-RFP at a MOI of ∼1-3. After infection, the cells were washed twice with media to remove unbound virus. Media containing the VSVΔG-RFP SARS-CoV-2 pseudotypes was harvested 28-30 hours after infection and clarified by centrifugation twice at 6000g then aliquoted and stored at −80 °C until used for antibody neutralization analysis.

#### Antibody neutralization assay using VSVΔG-RFP SARS-CoV-2

All sera were heat-inactivated for 30 minutes at 55 ⁰C prior to use in neutralization assay. Vero E6 cells stably expressing TMPRSS2 were seeded in 100 µl at 2.5×10^4^ cells/well in a 96 well collagen coated plate. The next day, 2-fold serially diluted serum samples were mixed with VSVΔG-RFP SARS-CoV-2 pseudotyped virus (100-300 focus forming units/well) and incubated for 1hr at 37 ⁰C. Also included in this mixture to neutralize any potential VSV-G carryover virus was 1E9F9, a mouse anti-VSV Indiana G, at a concentration of 600 ng/ml (Absolute Antibody). The serum-virus mixture was then used to replace the media on VeroE6 TMPRSS2 cells. 22 hours post infection, the cells were washed and fixed with 4% paraformaldehyde before visualization on an S6 FluoroSpot Analyzer (CTL, Shaker Heights OH). Individual infected foci were enumerated and the values compared to control wells without antibody. The focus reduction neutralization titer 50% (FRNT_50_) was measured as the greatest serum dilution at which focus count was reduced by at least 50% relative to control cells that were infected with pseudotyped virus in the absence of human serum. FRNT_50_ titers for each sample were measured in at least two technical replicates and were reported for each sample as the geometric mean.

#### Statistical analysis

GraphPad Prism software version 9 was used for generating dot plots, bar plots and the correlation images presented in this work. All statistical data analyses were carried out using R version 4.0.3. The departure of the data from a normal/Gaussian distribution was confirmed by the Shapiro-Wilk test and consequently, nonparametric, distribution-free tests were used for all comparisons throughout this work. Single comparisons between variables were performed using the two-tailed Mann-Whitney U test with continuity correction when the number of data points in each group was greater than seven. Else, the Wald–Wolfowitz runs test was employed to afford greater sensitivity to the analysis (Sprent, 2019). Univariate correlations involving continuous and categorical data were performed using the rank-based Spearman correlation analysis. The reported p-values are corrected for multiple hypothesis testing using the Benjamini-Hochberg procedure (Benjamini and Hochberg, 1995). Statistical significance for all comparisons was set at the critical values of p < 0.05 (*), p < 0.01 (**), p < 0.001 (***), and p < 0.0001 (****).

## SUPPLEMENTAL INFORMATION

**Supplementary Figure 1. GC B cell responses to SARS-CoV-2 mRNA vaccines are detectable in vaccine-draining ipsilateral lymph nodes.**

**(A)** Representative gating strategy for defining GC B cells (CD19^+^CD4^-^CD8^-^IgM^-^IgD^-^ CD38^+^CD27^lo/int^BCL6^+^), SARS-CoV-2-specific GC B cells (CD19^+^CD4^-^CD8^-^IgM^-^IgD^-^ CD38^+^CD27^lo/int^BCL6^+^HA^-^S^+^RBD^+/-^) and memory B cells (CD19^+^CD4^-^CD8^-^IgM^-^IgD^-^CD38^-^ CD27^+^).

**(B)** Quantification of GC B cells from ipsilateral lymph nodes of HDs, displayed as a percentage of total lymphocytes. Unpaired (orthogonal) changes between V2 and V3 are shown.

**(C)** Spearman correlation between HD age (years) and GC B cells (displayed as a percentage of lymphocytes) at V2 and V3.

**(D)** Orthogonal analysis of Full S^+^ RBD^-^ and Full S^+^ RBD^+^ GC B cells from HD ipsilateral lymph nodes, displayed as a percentage of GC B cells.

**(E and F)** Spearman correlation between HD age (years) and antigen-specific GC B cells (displayed as a percentage of GC B cells) at V2 and V3.

In (B and D), n=13 for V2 and V3. In (C, E and F), n =12 for V2 and V3. In (B-F), red data points = V2 and blue data points = V3. Statistical analysis: In (B and D), an unpaired Mann-Whitney U test with continuity correction was performed. In (C, E and F), correlations were determined using the Spearman’s *rho* with a 95% confidence interval. * P ≤ 0.05.

**Supplementary Figure 2. Tfh cell frequencies increase following immunization with SARS-CoV-2 mRNA vaccines.**

**(A)** Representative gating strategy for defining Tfh cells (CD4^+^CD8^-^CD19^-^CD45RA^-^CXCR5^hi^PD-1^hi^).

**(B)** Representative flow cytometry of Tfh cells in a putative quiescent cadaveric lymph node (LN Control, left) and a pre-pandemic tonsil sample (Tonsil Control, right).

**(C)** Orthogonal analysis of Tfh cells from HD ipsilateral lymph nodes at V2 and V3, displayed as a percentage of CD45RA^-^ CD4 T cells.

**(D)** (**Left**) Representative flow cytometry of activated ICOS^hi^PD-1^hi^ Tfh cells (CD4^+^CD8^-^CD19^-^ CD45RA^-^CXCR5^+^ICOS^hi^PD-1^hi^) from HD PBMC samples at V1, V2 and V3. (**Right**) Representative plot of CD38 expression, displayed as a histogram, in ICOS^hi^PD-1^hi^ Tfh cells.

**(E)** Spearman correlation between activated ICOS^hi^PD-1^hi^ Tfh cells, defined as in (D), from PBMCs (displayed as a percentage of CXCR5^+^ CD4 T cells) and antigen-specific GC B cells from ipsilateral lymph nodes (displayed as a percentage of lymphocytes) of HDs at V2 and V3.

In (C), n=13 for V2 and V3. In (E), n =13 for V2 and n = 11 for V3. In (C and E), red data points = V2 and blue data points = V3. Statistical analysis: In (C), an unpaired Mann-Whitney U test with continuity correction was performed. In (E), correlations were determined using the Spearman’s *rho* with a 95% confidence interval. * P ≤ 0.05.

**Supplementary Figure 3. SARS-CoV-2 mRNA vaccines induce the generation of antigen-specific memory B cells.**

**(A)** Orthogonal analysis of SARS-CoV-2-specific memory B cells from HD ipsilateral lymph nodes (LNs) at V2 and V3, displayed as a percentage of lymphocytes.

**(B)** Orthogonal analysis of SARS-CoV-2-specific memory B cells from HD PBMCs at V2 and V3, displayed as percentage of lymphocytes.

**(C)** Spearman correlation between SARS-CoV-2-specific memory B cells from ipsilateral lymph nodes (LN) and PBMCs of HDs, both displayed as a percentage of lymphocytes.

**(D)** Orthogonal analysis of plasmablasts from PBMCs of HDs at V2 and V3, displayed as a percentage of lymphocytes.

**(E)** Spearman correlation between plasmablasts from HD ipsilateral lymph nodes (LN) and PBMCs at V2 and V3, both represented as a percentage of lymphocytes.

In (A and B), n=13 for V2 and V3. In (C - E), n =13 for V2 and n = 11 for V3. In (A-E), red data points = V2 and blue data points = V3. Statistical analysis: In (A, B, and D), an unpaired Mann-Whitney U test with continuity correction was performed. In (C and E), correlations were determined using the Spearman’s *rho* with a 95% confidence interval. * P ≤ 0.05, ** P ≤ 0.01, *** P ≤ 0.001.

**Supplementary Figure 4. Kidney transplant recipients have blunted germinal center responses which correlates with reduced B cell and humoral responses.**

**(A)** Quantification of the total cell yield from ipsilateral lymph nodes (LNs) of HDs and KTX recipients at V2 and V3.

**(B)** Quantification of SARS-CoV-2-specific memory B cells in PBMCs from HDs and KTX recipients, displayed as a percentage of B cells.

**(C)** Quantification of SARS-CoV-2-specific memory B cells from ipsilateral lymph nodes (LN) of HDs and KTX recipients, displayed as a percentage of memory B cells.

**(D)** Spearman correlation between SARS-CoV-2-specific memory B cells (from PBMCs) and SARS-CoV-2-specific GC B cells (from ipsilateral lymph nodes, LN) from HDs and KTX recipients at V2 and V3, both displayed as a percentage of B cells.

**(E)** Quantification of plasmablasts in PBMCs from HDs and KTX recipients, displayed as a percentage of B cells.

**(F and G)** Spearman correlation between SARS-CoV-2 binding (F) and neutralizing (G) antibodies (against the D614G mutant) and SARS-CoV-2-specific GC B cells (from ipsilateral lymph nodes, displayed as a percentage of B cells) from HDs and KTX recipients at V2 and V3.

**(H)** Spearman correlation between neutralizing antibody titers against the D614G mutant and activated ICOS^hi^PD-1^hi^ Tfh cells from ipsilateral lymph nodes (top) or PBMCs (bottom), shown as a percentage of CXCR5^+^ CD4 T cells.

In (A), for HD: n = 13 for V2 and V3; for KTX: n = 5 for V2 and n = 6 for V3. In (B and E), for HD: n =13 for V2 and n = 11 for V3; for KTX: n = 2 for V2 and n = 8 for V3. In (C and D), for HD: n = 13 for V2 and V3; for KTX: n = 3 for V2 and n = 7 for V3. In (F-H), for HD: n = 12 for V2 and V3; for KTX: n = 2 for V2 and n = 7 for V3. In (A-H), a circle is used to represent HDs, a triangle is used to represent KTX recipients, and a square is used to indicate a KTX recipient with a prior SARS-CoV-2 infection; red data points = V2 and blue data points = V3. Statistical analysis: In (A), an unpaired Mann-Whitney U test was performed. In (B, C and E), the Wald-Wolfowitz runs test was performed. In (D and F-H), correlations were determined using the Spearman’s *rho* with a 95% confidence interval. * P ≤ 0.05, ** P ≤ 0.01, *** P ≤ 0.001, **** P ≤ 0.0001.

**Supplementary Figure 5. SARS-CoV-2-specific CD4 and CD8 T cells are reduced in KTX PBMCs**

**(A)** Representative flow cytometry gating strategy for AIM assays on PBMC samples.

**(B)** Quantification of AIM^+^ CD4 T cells as a percentage of total CD4 T cells. Paired (longitudinal, left) or unpaired (orthogonal, right) analyses of PBMC samples from HDs and KTX recipients at V1, V2 and V3 are shown.

**(C and D)** Quantification of AIM^+^ CD4 T cell subsets (C) and AIM^+^ CXCR5^+^ CD4 T cell subsets (D) in PBMCs from KTX recipients, shown as a frequency of AIM^+^ CD45RA^-^ CD4 T cells.

**(E)** Quantification of AIM^+^ CD8 T cells (IFN-γ^+^ 41BB^+^), shown as a percentage of total CD8 T cells, in PBMC samples from HDs and KTX recipients.

**(F)** Quantification of AIM^+^ CD8 T cells (cells expressing at least 3 of 5 activation markers: CD107a, 41BB, CD200, CD40L, and IFN-γ), shown as a percentage of total CD8 T cells, in PBMC samples from HDs and KTX recipients.

**(G and H)** Quantification of total CD45RA^-^ CD4 T cell subsets in PBMC samples from HDs (G) and KTX recipients (H), shown as a frequency of total CD45RA^-^ CD4 T cells.

In (B-H), for HD: n = 11 for V1and V3, n = 9 for V2; for KTX: n = 5 for V1, n = 6 for V2, and n = 7 for V3. In (B and E-F), a circle is used to represent HDs and a triangle is used to represent KTX; white data points = V1, red data points = V2 and blue data points = V3. Statistical analysis: In (B-H), the Wald-Wolfowitz runs test was used to perform an exact comparison between the two data distributions of interest. * P ≤ 0.05, ** P ≤ 0.01, *** P ≤ 0.001, **** P ≤ 0.0001.

## Notes

### Author Declarations

Written informed consent for participation was obtained according to the Declaration of Helsinki and protocols were approved by the Institutional Review Board of the University of Pennsylvania.

