## Supplementary figures and images for "Germinal center responses to SARS-CoV-2 mRNA vaccines in healthy and immunocompromised individuals"

### Supplemental figures

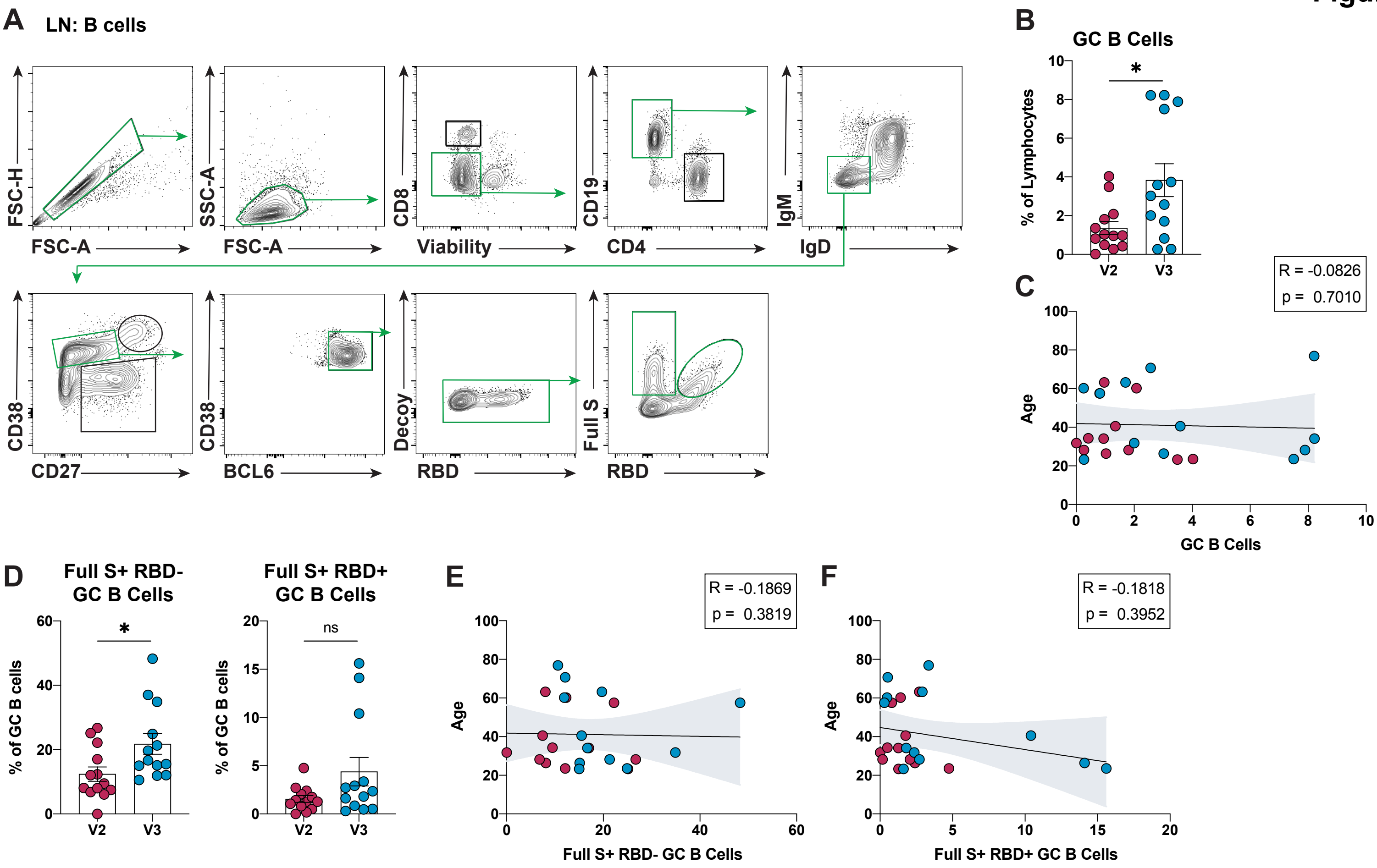

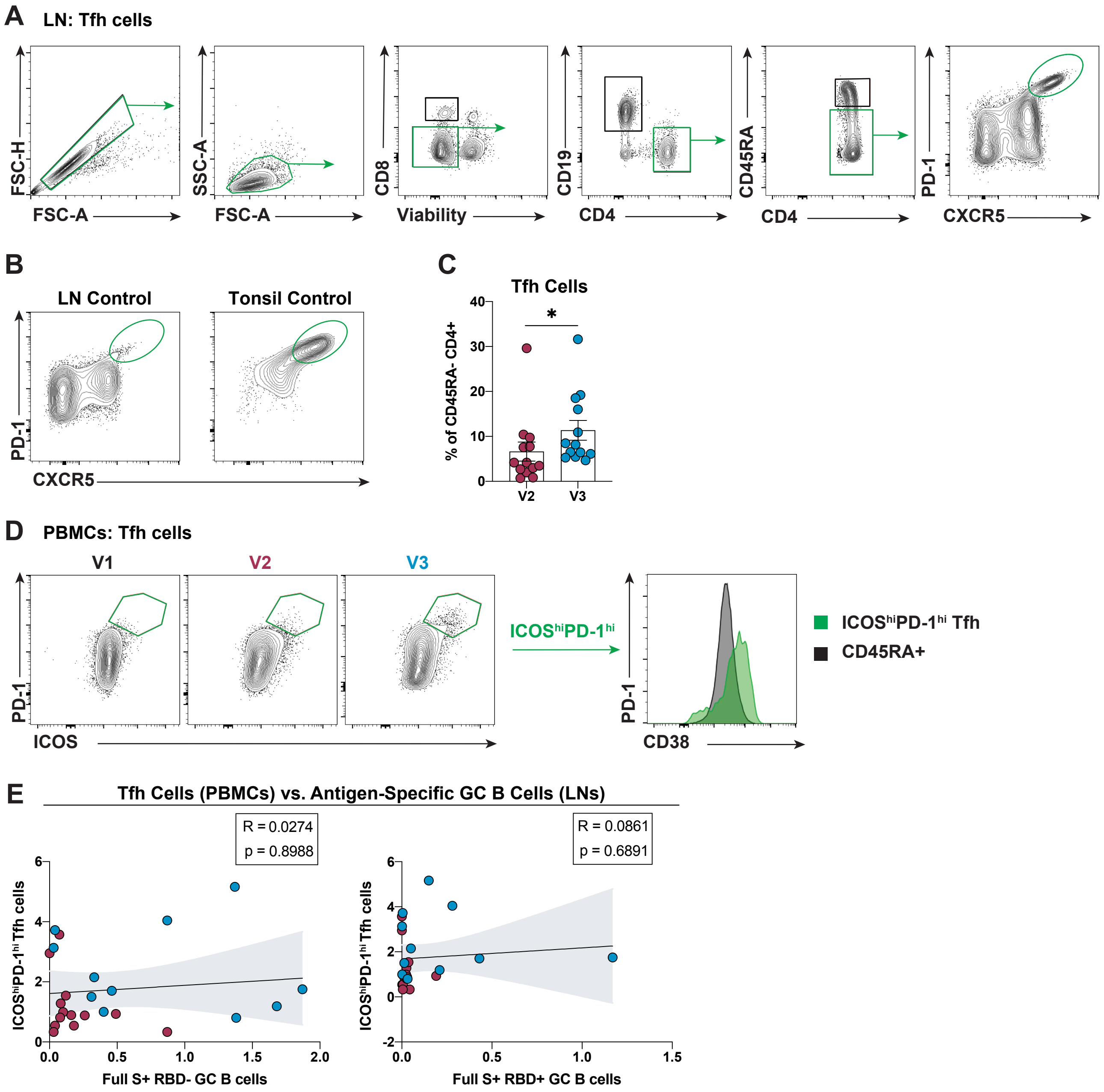

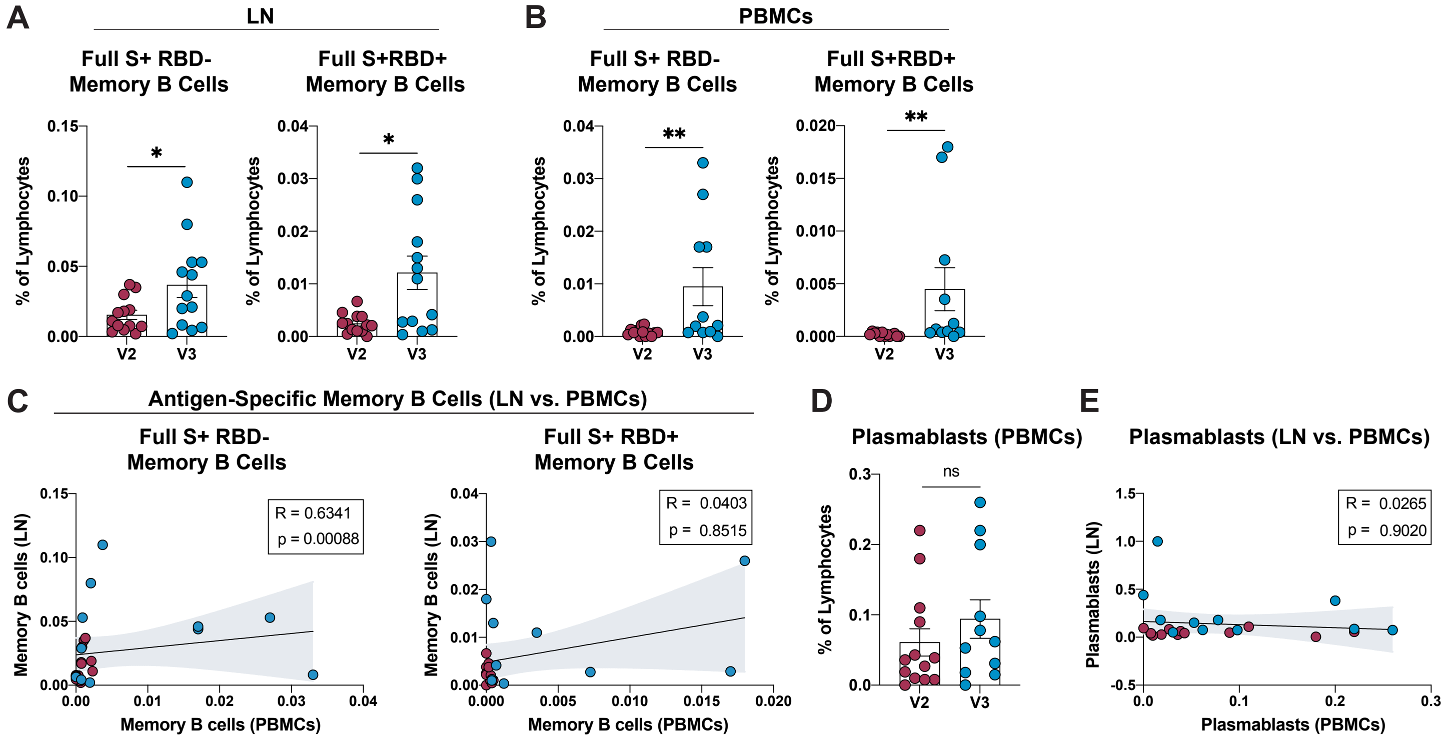

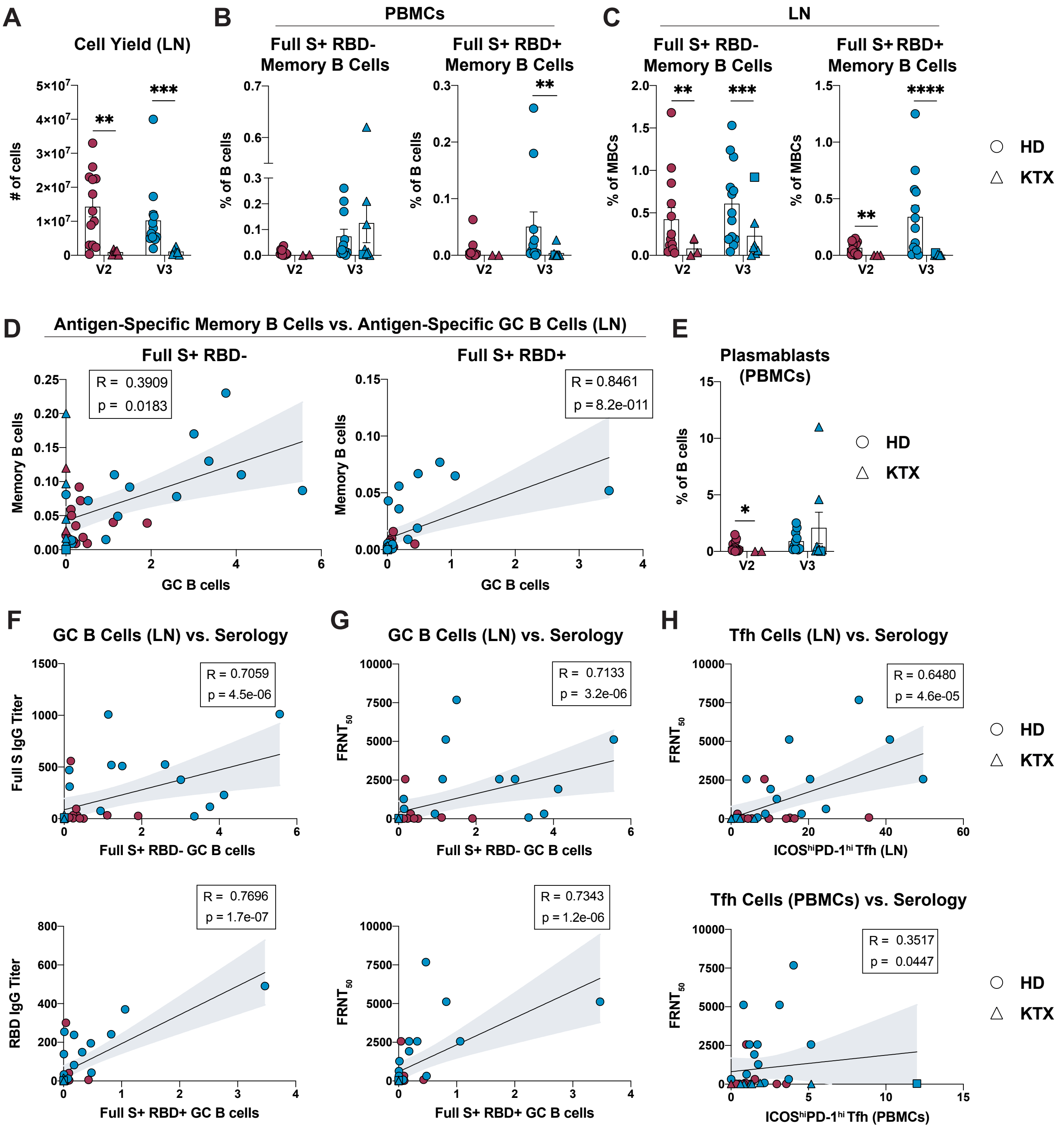

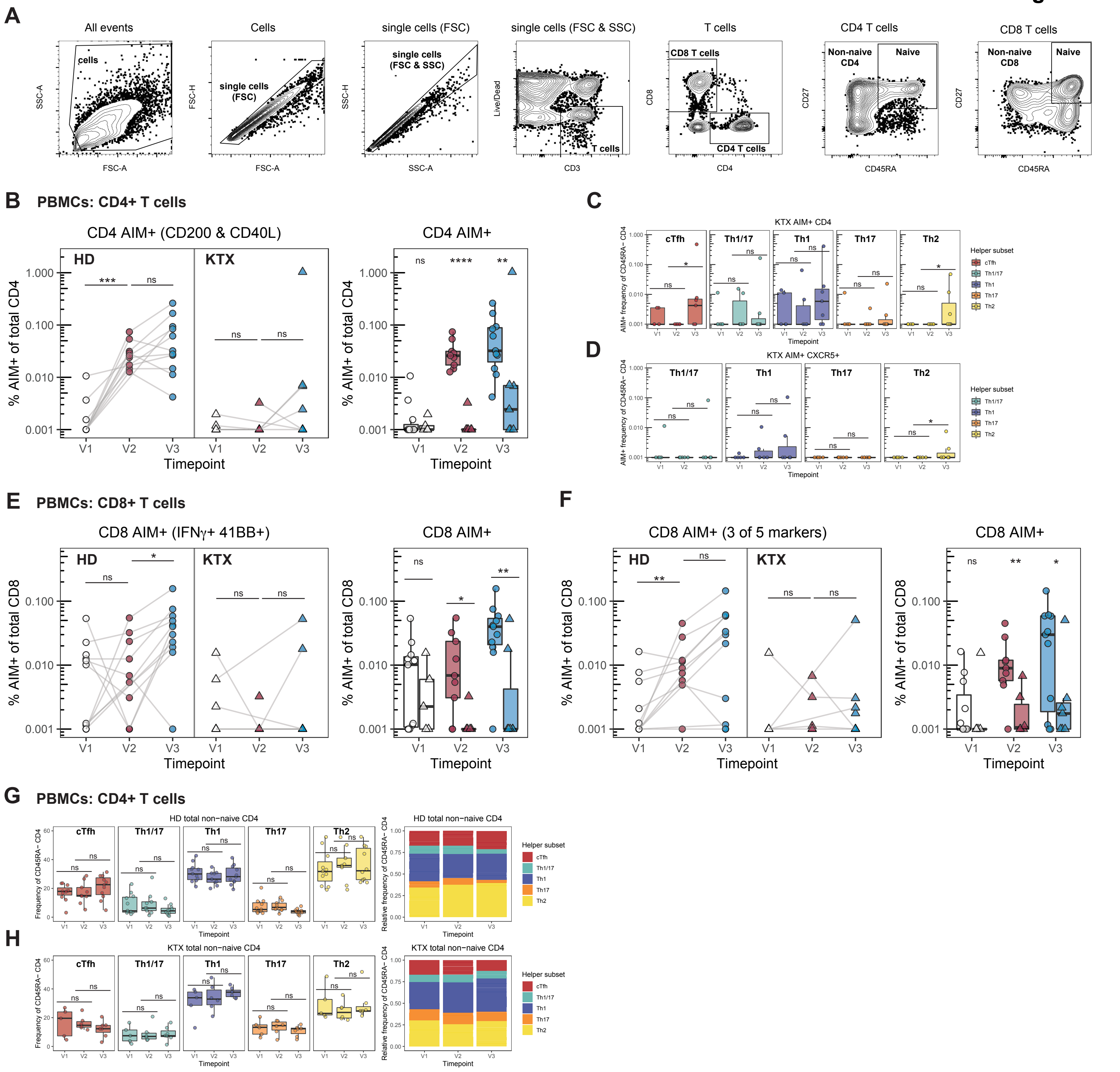
