## Supplementary material for "Germinal center responses to SARS-CoV-2 mRNA vaccines in healthy and immunocompromised individuals": Tables

**Table 1. Lymph node FNA cohort demographics and baseline clinical characteristics**

| | **Healthy** | **Kidney Transplant Recipients** |
|---|---|---|
| **Variable** | Total N=15 | Total N=10 |
| | N (%) | N (%) |
| **Age (Median [Range])** | 34 [23-76] | 52 [28-66] |
| **Sex** | | |
| Males | 6 (40) | 7 (70) |
| Females | 9 (60) | 3 (30) |
| **Race** | | |
| African American | 0 (0) | 5 (50) |
| Asian | 5 (33.3) | 1 (10) |
| White | 10 (66.7) | 4 (40) |
| Other | 0 (0) | 0 (0) |
| **Ethnicity** | | |
| Not of Hispanic, Latinx, or Spanish origin | 14 (93.3) | 9 (90) |
| Hispanic, Latinx, or Spanish origin | 1 (6.7) | 1 (10) |
| **Etiology of Renal Failure** | | T2DM (4), HTN (1), PCKD (3), FSGS (1), Lupus (1) |
| **Duration of Dialysis (Months), (Median [Range])** | | 49.3 [0-107.5] |
| **Time since Kidney Transplant (Months), (Median [Range])[a]** | | 12.6 [-0.3 - 63.1] |
| **Type of donor organ** | | DDKT (8), LUKT (2) |
| **Number of previous transplants (Mean [Range])** | | 0 [0-0] |
| **cPRA (%) (Median [Range])** | | 0 [0-39] |
| **eGFR (mL/min/1.73m^2), (Median [Range])[a, b]** | | 51.5 [10 - >60] |
| **WBC (tho/uL), (Median [Range])** | | 5.9 [3.4-11.1] |
| **ALC (tho/uL), (Median [Range])** | | 0.5 [0-1.6] |
| **Hgb (g/dL), (Median [Range])** | | 11.6 [8.7-15.8] |
| **Creatinine (mg/dL), (Median [Range])** | | 1.63 [0.93-8.74] |
| **Immunosuppression** | | |
| Thymoglobulin <1 yr. Prior ("n", Median [Range in days])[a] | | 6, 57 [-11 - 158] |

| | | |
|---|---|---|
| Thymoglobulin >1 yr. Prior ("n", Median [Range in days]) | | 4, 1118 [1156-1883] |
| Maintenance | | |
| Tacrolimus, Mycophenolate mofetil, Prednisone | | 8 |
| Cyclosporine, Prednisone | | 1 |
| **History of SARS-CoV2 infection** | 0 (0) | 1 (10) |
| **Vaccine** | BNT162b2 (10), mRNA-1273 (5) | BNT162b2 (9), mRNA-1273 (1) |

[a] One patient received the 1st dose of vaccination 9 days prior to transplant.

[b] A value of 61 was used for any eGFR >60 for calculation of the median.

T2DM - Type 2 diabetes mellitus, HTN - hypertension, PCKD - polycystic kidney disease, FSGS - focal segmental glomerular sclerosis, DDKT - Deceased donor kidney transplant, LURT - Living unrelated kidney transplant, cPRA – Calculated panel-reactive antibody, eGFR – Estimated glomerular filtration rate, WBC – White blood cell count, ALC – Absolute lymphocyte count, Hgb - Hemoglobin.

**Table 2. T cell AIM cohort demographics and baseline clinical characteristics**

| | **Healthy** | **Kidney Transplant Recipients** |
|---|---|---|
| **Variable** | Total N=11 | Total N=10 |
| | N (%) | N (%) |
| **Age (Median [Range])** | 33 [23-62] | 56 [28-70] |
| **Sex** | | |
| Males | 3 (27.3) | 4 (40) |
| Females | 8 (72.7) | 6 (60) |
| **Race** | | |
| African American | 0 (0) | 7 (70) |
| Asian | 2 (18.2) | 1 (10) |
| White | 9 (81.8) | 2 (20) |
| Other | 0 (0) | 0 (0) |
| **Ethnicity** | | |
| Not of Hispanic, Latinx, or Spanish origin | 10 (90.9) | 10 (100) |
| Hispanic, Latinx, or Spanish origin | 1 (9.1) | 0 (0) |
| **Etiology of Renal Failure** | | T2DM (3), HTN (2), T2DM/HTN (1), PCKD (3), FSGS (1) |
| **Duration of Dialysis (Months), (Median [Range])** | | 49.3 [0-107.5] |
| **Time since Kidney Transplant (Months), (Median [Range])[a]** | | 12.6 [1.1 - 63.1] |
| **Type of donor organ** | | DDKT (8), LUKT (2) |
| **Number of previous transplants (Mean [Range])** | | 0 [0-0] |
| **cPRA (%) (Median [Range])** | | 0 [0-39] |
| **eGFR (mL/min/1.73m^2), (Median [Range])[a, b]** | | 48 [35 - >60] |
| **WBC (tho/uL), (Median [Range])** | | 5.9 [2.8-11.1] |
| **ALC (tho/uL), (Median [Range])** | | 0.6 [0.2-2.3] |
| **Hgb (g/dL), (Median [Range])** | | 11.6 [8.7-13.4] |
| **Creatinine (mg/dL), (Median [Range])** | | 1.52 [0.8-2.05] |
| **Immunosuppression** | | |

| | | |
|---|---|---|
| Thymoglobulin <1 yr. Prior ("n", Median [Range in days])[a] | | 5, 62 [31 - 158] |
| Thymoglobulin >1 yr. Prior ("n", Median [Range in days]) | | 5, 1270 [1118-1883] |
| Maintenance | | |
| Tacrolimus, Mycophenolate mofetil, Prednisone | | 7 |
| Cyclosporine, Prednisone | | 3 |
| **History of SARS-CoV2 infection** | 0 (0) | 0 (0) |
| **Vaccine** | BNT162b2 (8), mRNA-1273 (3) | BNT162b2 (9), mRNA-1273 (1) |

[a] A value of 61 was used for any eGFR >60 for calculation of the median. T2DM - Type 2 diabetes mellitus, HTN - hypertension, PCKD - polycystic kidney disease, FSGS - focal segmental glomerular sclerosis, DDKT - Deceased donor kidney transplant, LURT - Living unrelated kidney transplant, cPRA – Calculated panel-reactive antibody, eGFR – Estimated glomerular filtration rate, WBC – White blood cell count, ALC – Absolute lymphocyte count, Hgb - Hemoglobin.

**Table 3. Flow Cytometry Panel for FNA Analysis, related to Figures 1-5 and Figures S1-4.**

| Antibody/Conjugate | Conjugation | Dilution Factor | Clone |
|---|---|---|---|
| CXCR5 | BUV395 | 1:40 | RF8B2 |
| IgM | BUV496 | 1:200 | UCH-B1 |
| CD4 | BUV563 | 1:200 | SK3 |
| CD11c | BUV661 | 1:100 | B-ly6 |
| IgD | BUV737 | 1:200 | IA6-2 |
| CD45RA | BUV805 | 1:200 | HI100 |
| Full S Probe | BV421 | 1:200 | N/A |
| CCR6 | BV480 | 1:33 | 11A9 |
| CXCR3 | BV510 | 1:20 | G025H7 |
| CD138 | BV605 | 1:50 | MI15 |
| ICOS | BV650 | 1:50 | C398.4A |
| CD20 | BV750 | 1:200 | 2H7 |
| CD38 | BV785 | 1:100 | III 155 |
| HA Probe | Alexa Fluor 488 | 1:200 | N/A |
| CD8 | PerCP-eFluor 710 | 1:200 | OKT-8 |
| RBD Probe | PE | 1:200 | N/A |
| CCR4 | PE-CF594 | 1:20 | 1G1 |
| CD27 | PE-Cy5 | 1:50 | O323 |
| CD19 | PE-Cy5.5 | 1:200 | SJ25C1 |
| PD-1 | PE-Cy7 | 1:50 | EH12.2H7 |
| Bcl6 | Alexa Fluor 647 | 1:50 | K112-91 |
| CD11b | Alexa Fluor 700 | 1:100 | ICRF44 |
| Live/Dead | eFluor 780 | 1:2000 | N/A |

**Table 4. Flow Cytometry Panel for Activation Induced Marker (AIM) analysis, related to Figure 6 and Figure S5.**

| Antibody/Conjugate | Conjugation | Dilution Factor | Clone |
|---|---|---|---|
| CD4 | BUV395 | 1:400 | SK3 |
| CD8 | BUV496 | 1:400 | RPA-T8 |
| CD45RA | BUV615 | 1:2000 | HI100 |
| CD27 | BUV737 | 1:400 | L128 |
| CD3 | BUV805 | 1:800 | UCHT1 |
| CXCR3 | BV421 | 1:800 | G02587 |
| CCR7 | BV650 | 1:400 | G043H7 |
| CD69 | BV605 | 1:400 | FN50 |
| CD40L | BV711 | 1:50 | 24-31 |
| CD107a | BV785 | 1:100 | H4A3 |
| IFNγ | FITC | 1:400 | 4S.B3 |
| CD200 | PE | 1:100 | A18042B (OX2) |
| OX40 | PE-Cy7 | 1:1600 | Ber-ACT35 |
| 41BB | AF647 | 1:400 | 4B4-1 |
| CXCR5 | APC R700 | 1:100 | RF8B2 |
| CCR6 | APC-Cy7 | 1:800 | G034E3 |
| Live/Dead | Ghost Dye Violet 510 | 1:800 | N/A |
